# An integrative multi-omics approach in Sjögren’s Syndrome identifies novel genetic drivers with regulatory function and disease-specificity

**DOI:** 10.1101/2020.09.14.20192211

**Authors:** María Teruel, Guillermo Barturen, Manuel Martínez-Bueno, Miguel Barroso, Olivia Castelli, Elena Povedano, Martin Kerick, Francesc Català-Moll, Zuzanna Makowska, Anne Buttgereit, PRECISESADS Clinical Consortium, PRECISESADS Flow Cytometry Study Group, Jacques-Olivier Pers, Concepción Marañón, Esteban Ballestar, Javier Martin, Elena Carnero-Montoro, Marta E. Alarcón-Riquelme

**Affiliations:** GENYO, Center for Genomics and Oncological Research Pfizer/University of Granada/Andalusian Regional Government, 18016 Granada, Spain; IBPLN- CSIC, Instituto de Parasitología y Biomedicina López-Neyra, Consejo Superior de Investigaciones Científicas, Granada 18016, Spain; IDIBELL, Bellvitge Biomedical Research Institute 08907 L’Hospitalet de Llobregat, Barcelona, Spain; Epigenetics and Immune Disease Group, Josep Carreras Research Institute (IJC), 08916 Badalona, Barcelona, Spain; Pharmaceuticals Division, Bayer Pharma Aktiengesellschaft, Berlin, Germany; Université de Brest, INSERM, Labex IGO, CHU de Brest, Brest, France; Institute for Environmental Medicine, Karolinska Institutet, 171 67 Solna, Sweden

**Keywords:** Sjögren’s Syndrome, DNA methylation, epigenome-wide association study, RNAseq, meQTL, eQTL, omics, gene-by-environment interaction

## Abstract

Primary Sjögren’s syndrome (SS) is a systemic autoimmune disease characterized by lymphocytic infiltration and damage of exocrine salivary and lacrimal glands. The etiology of SS is complex with environmental triggers and genetic factors involved. By conducting an integrated multi-omics study we identified vast coordinated hypomethylation and overexpression effects, that also exhibit increased variability, in many already known IFN-regulated genes. We report a novel epigenetic signature characterized by increased DNA methylation levels in a large number of novel genes enriched in pathways such as collagen metabolism and extracellular matrix organization. We identified new genetic variants associated with SS that mediate their risk by altering DNA methylation or gene expression patterns, as well as disease-interacting genetic variants that exhibit regulatory function only in the SS population. Our study sheds new light on the interaction between genetics, DNA methylation, gene expression and SS, and contributes to elucidate the genetic architecture of gene regulation in an autoimmune population.

## INTRODUCTION

Primary Sjögren’s syndrome (SS) [MIM 270150] is a systemic autoimmune disease characterized by lymphoid infiltration and tissue damage of the exocrine glands, mainly the salivary and lacrimal glands ^1^. The prevalence of SS is about 1% of the World population being the main risk group being middle-aged women. SS patients show great clinical heterogeneity that is manifested at the serological, genetic, and cellular level and in their capacity to respond to treatment. SS patients with anti-Ro/SSA or anti-La/SSB autoantibodies usually develop a more severe disease with systemic manifestations and the appearance of lymphomas in a percentage of them ^2^.

The etiology of SS is complex and not completely understood; environmental, genetic and epigenetic factors are known to be involved in its development^1,3–5^. Genome-wide association studies (GWAS) have signified an important advance for understanding the mechanisms involved in the pathogenesis of SS. There are currently around 10 well-known risk *loci* implicated in SS susceptibility that have been revealed by GWAS studies^3–5^. The *loci* identified suggest the importance of the adaptive and innate immune responses in SS pathology, especially the interferon (IFN) signaling pathway. Other important pathways include B cells signaling and autoantibody production, the NF-kB signaling pathway and the T cell activation through the major histocompatibility complex ^6^. Many of the variants detected in GWAS studies are located in non-coding intron or intergenic regions, suggesting that they have a regulatory role that is fairly unexplored ^7,8^. Moreover, the identified SS-associated genetic variants can only explain a small proportion of the heritability observed in SS^9^, suggesting the possible contribution of many gene variants at the low-frequency spectrum, with lower effect sizes, and/or the implication of gene-by-environment interactions.

Functional approaches based on genome-wide data of the epigenome or the transcriptome are allowing to dive depth into the genetics of transcriptional regulation by detecting variants with potential regulatory effect, i.e., expression quantitative trait loci (eQTLs)^8^ or methylation quantitative trait loci (meQTLs) discovery^10^. An increasing number of studies are showing widespread regulatory effects of disease associated genetic variants and are helping to mechanistically explain the genetic risk of disease. Functional genomics approaches have been applied very successfully in cancer research, and to some extent to autoimmune diseases as well, especially in rheumatoid arthritis (RA)^11,12^ and systemic lupus erythematosus (SLE)^13,14^, but only recently in SS^15^.

Epigenetic alterations can be integrators of the complex interaction between genes and the environment and are known to play a relevant role in autoimmunity by altering gene expression profiles in response to genetic factors, changing environment and immunological conditions^16,17^. Recently, a few epigenome-wide association studies (EWAS) have been performed in different cell types and have consistently found a pervasive hypomethylation in CpG sites within genes related with the type I IFN signaling, confirming this pathway as key in SS pathology ^18–20^. Likewise, gene expression profiling of minor salivary glands or peripheral blood showed a consistent upregulation of IFN-inducible genes associated with SS, which was found most pronounced in the subset of cases serologically defined by increased titers of anti-Ro/SSA and anti-La/SSB autoantibodies ^21,22^.

Despite an increasing number of studies interrogating different layers of molecular information, the existing integrative approach for SS mainly focuses on deciphering the regulatory roles of previously known disease associated genetic variants discovered by GWAS ^15,23^. Therefore, these studies have not contributed to the identification of new risk variants associated with SS with regulatory function that could explain further the missing heritability of this complex disease ^20^. Furthermore, the vast majority of large-scale QTL studies have been performed in healthy populations, and do not directly compare the genetic regulatory effects between cases and controls, which could very much be different as the pathological condition imposes a different environmental and cellular context in affected individuals, especially in immune-related conditions. In this regard, previous experimental studies have shown that eQTL and meQTL effects are highly context-specific, as for example cell-type specific effects have been found when studying different blood cell types^24–26^, and experimental studies of ex-vivo activation of immune cells have shown that eQTLs profiles are altered upon different stimuli^27,28^.

This work represents the first epigenome- and transcriptional-wide integrative study that combines high throughput data on genetics, DNA methylation and gene expression in a large group of SS patients and healthy subjects. It contributes, on the one hand, to identify new molecular pathways involved in SS. On the other hand, this work identifies important novel genetic variants implicated in the disease through changes in DNA methylation or gene expression variants with regulatory effects exhibiting disease-specificity, therefore contributing to elucidate the genetic architecture of gene regulation in an autoimmune population.

## RESULTS

An overview of the methodology used and main analyses performed in this study is illustrated in **Fig 1**.

**Figure 1.**
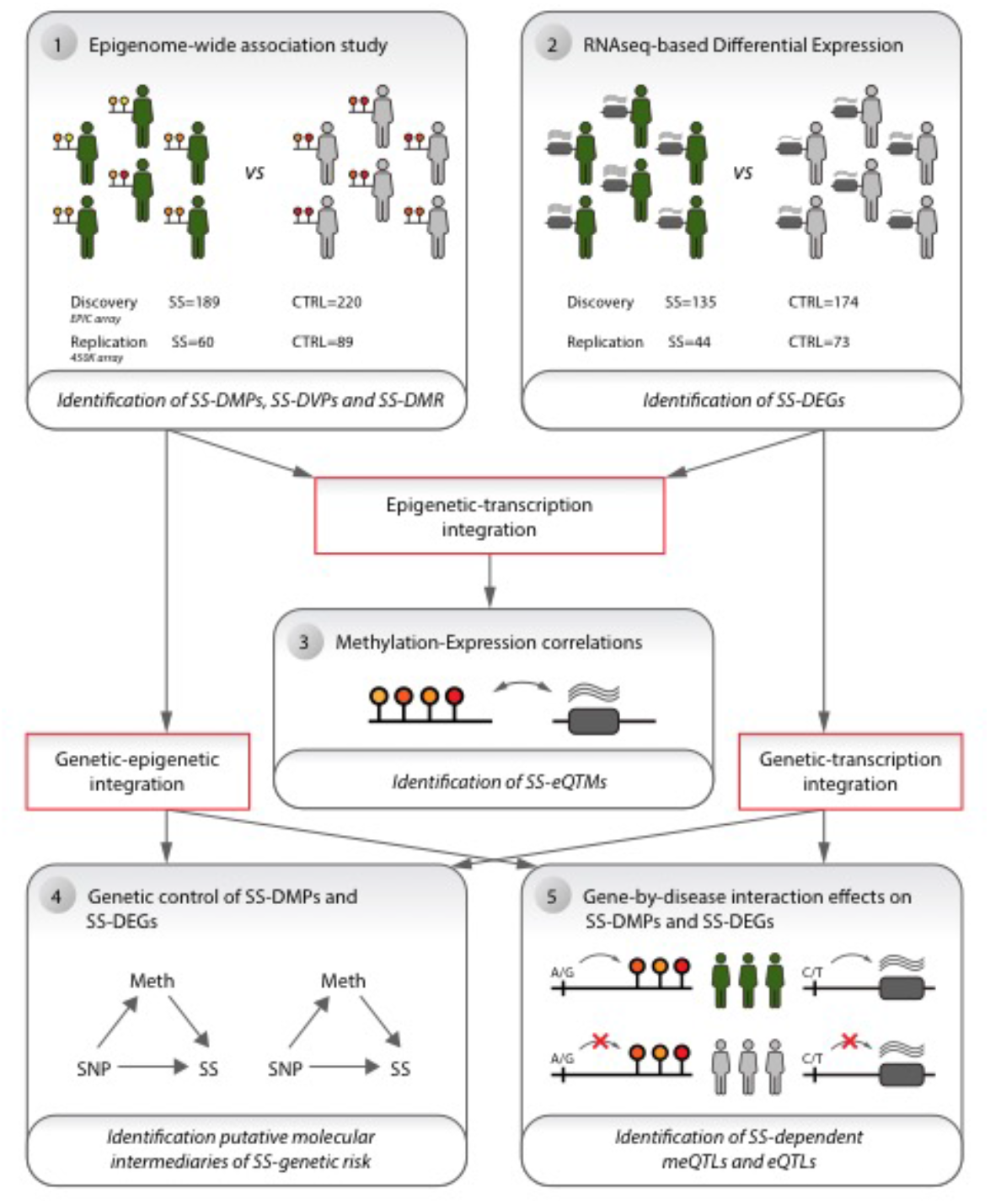
Study design overview. An epigenome-wide association study was first performed (**1**) to find differentially methylated positions (DMPs), differentially methylated regions (DMRs) and variable methylated positions (VMPs) associated with SS. This was followed by differential expression analyses based on RNA-seq data to detect differentially expressed genes (DEGs) (**2**). Thirdly, epigenetic and gene expression data was integrated by performing expression quantitative trait methylation (eQTMs) analysis to explore the correlation between SS-DMRs and SS-DEFs and to identify eQTMs associated with SS (SS-eQTMs) (**3**). Then, epigenetic and gene expression data was integrated with genetics to find genetic variants that could mediate their risk contribution to disease by altering DNA methylation or gene expression landscapes (**4**). Finally, genetic variants with regulatory effects that influence DMPs and DEGs were discovered only in the context of SS disease by performing gene-by-disease interaction meQTL and eQTL analyses (**5**).

### Differentially methylated positions associated with SS

We explored the DNA methylation patterns associated with SS comparing the whole blood DNA methylation level between 189 SS patients and 220 heathy subjects (**Supplementary Table 1**) using the Infinium MethylationEPIC BeadChip with which we could interrogate 776,284 autosomic CpG sites. In total, we observed 118 differential methylated positions (DMPs) associated with SS at a Bonferroni-corrected threshold of P< 6.4 x 10^-08^ (**Supplementary Table 2)**. The majority of SS-associated DMPs exhibited decreased methylation levels in SS patients compared with controls (91.5%), supporting the overall hypomethylation previously described in SS patients **(Fig. 2A)** ^18,19^. The 118 SS-associated DMPs were annotated to 52 unique genes and 7 intergenic regions. The majority of them fell within promoters (49.2 %) and gene bodies (40.7%), and only 4.2 % were located in the 3’UTR. SS-associated DMPs were mainly distributed in open sea and shore CpGs and only 5 DMPs fell in CpG islands (**Supplementary Fig. 1**). We included an independent cohort formed by 60 SS patients and 89 healthy individuals (**Supplementary Table 1**) from whom we had DNA methylation data available from the 450K array which contained half of the SS-associated DMPs detected in our discovery cohort. At a P< 0.05 we could successfully replicate 84.7% (**Supplementary Table 2**). The top 10 SS-associated DMPs with an average methylation difference |Δβ| > 0.1 were located within *IFI44L, MX1, PARP9-DTX3L, NLRC5, IFIT1, IFIT3, IFITM1, PLSCR1, PDE7A* and *DDX60* genes, all of them regulated by IFN levels (see **Table 1A)**. The most significant DMPs was the cg05696877 probe in the *IFI44L* gene for which an average methylation difference |Δβ| = 0.36 was observed (P = 3.5 x 10^-27^; FDR = 2.7 x10^-21^) between SS and controls (**Supplementary Fig. 2a)**. We searched for CpGs that exhibit differences in DNA methylation variability between SS patients and controls and named these variable methylated positions (VMPs) **(Supplementary Table 2)**. We observed that 80% of SS-associated DMPs also exhibited evidence of increased DNA methylation variability at a significance level of P < 0.05 in SS patients compared with healthy controls. In our replication cohort, we could replicate 83.3 % of the VMPs observed. The increased DNA methylation variability associated with SS is especially pronounced for *MX1 and PARP9-DXT3L* genes for which we observed the largest variability differences (**Supplementary Fig. 2b and 2c**). Functional analyses for genes underlying SS-DMPs confirmed an enrichment of GO terms related with IFN signaling (GO:0060337; P= 3.5×10^-31^; GO:0034340; P= 31.37 ×10^-30^) as well as with defense response to virus (GO:0051607; P= 2.3 ×10^-23^) and cytokine-mediated signaling (GO:0019221; P=1.63 ×10^-19^) (**Supplementary Table 4**).

**Table 1.** Top differentially methylated positions and differentially expressed genes associated with Sjögren’s syndrome (SS-DMPs and SS-DEGs, respectively)

| **A. Top Differentially Methylated Positions associated with SS (SS-DMPs)** |  |  |  |  |  |  |
| --- | --- | --- | --- | --- | --- | --- |
| CpG site | Gene | Position<br>(hg38 Chr:bp) | Discovery cohort |  | Replication cohort |  |
| | | | $\beta$ | p-value | $\beta$ | p-value |
| cg05696877 | *IFI44L* | 1: 78623085 | -0.34 | $3.5 \times 10^{-27}$ | -0.24 | $2.3 \times 10^{-05}$ |
| cg21549285 | *MX1* | 21: 41427165 | -0.25 | $5.5 \times 10^{-19}$ | -0.25 | $5.4 \times 10^{-05}$ |
| cg22930808 | *PARP9\|DTX3L* | 3: 122562985 | -0.21 | $2.9 \times 10^{-18}$ | -0.23 | $2.1 \times 10^{-05}$ |
| cg07839457 | *NLRC5* | 16: 56989061 | -0.18 | $7.6 \times 10^{-20}$ | -0.14 | $6.0 \times 10^{-04}$ |
| cg05552874 | *IFIT1* | 10: 89393337 | -0.15 | $1.1 \times 10^{-16}$ | -0.14 | $2.2 \times 10^{-04}$ |
| cg06188083 | *IFIT3* | 10: 89333249 | -0.15 | $8.9 \times 10^{-18}$ | -0.14 | $1.9 \times 10^{-04}$ |
| cg23570810 | *IFITM1* | 11: 315103 | -0.14 | $1.5 \times 10^{-15}$ | -0.1 | $2.7 \times 10^{-03}$ |
| cg06981309 | *PLSCR1* | 3: 146543168 | -0.12 | $6.8 \times 10^{-18}$ | -0.1 | $1.0 \times 10^{-03}$ |
| cg14864167 | *PDE7A* | 8: 65838898 | -0.13 | $2.5 \times 10^{-10}$ | -0.1 | $1.1 \times 10^{-02}$ |
| cg05883128 | *DDX60* | 4: 168317931 | -0.1 | $1.2 \times 10^{-14}$ | -0.09 | $8.5 \times 10^{-04}$ |
| **B. Top Differentially Expressed genes associated with SS (SS-DEGs)** |  |  |  |  |  |  |
| Ensembl.ID | gene | Position<br>(hg38 Chr:bp TSS) | Discovery cohort |  | Replication cohort |  |
|  |  |  | log2FC | p-value | log2FC | p-value |
| ENSG00000137959 | *IFI44L* | 1:78620448 | 3.12 | $7.6 \times 10^{-55}$ | 2.89 | $1.6 \times 10^{-19}$ |
| ENSG00000134321 | *RSAD2* | 2:6877777 | 3.12 | $3.9 \times 10^{-50}$ | 2.98 | $4.7 \times 10^{-18}$ |
| ENSG00000184979 | *USP18* | 22:18150170 | 2.36 | $3.8 \times 10^{-48}$ | 2.3 | $5.9 \times 10^{-20}$ |
| ENSG00000137965 | *IFI44* | 1:78649831 | 2.25 | $4.5 \times 10^{-48}$ | 2.13 | $8.4 \times 10^{-17}$ |
| ENSG00000111331 | *OAS3* | 12:112938474 | 2.12 | $2.0 \times 10^{-46}$ | 2.13 | $4.6 \times 10^{-18}$ |
| ENSG00000088827 | *SIGLEC1* | 20:3686970 | 2.61 | $5.6 \times 10^{-45}$ | 2.71 | $2.0 \times 10^{-21}$ |
| ENSG00000115155 | *OTOF* | 2:26457203 | 3.55 | $6.4 \times 10^{-45}$ | 3.46 | $7.8 \times 10^{-21}$ |
| ENSG00000134326 | *CMPK2* | 2:6850650 | 2.3 | $8.4 \times 10^{-45}$ | 2.16 | $1.8 \times 10^{-16}$ |
| ENSG00000089127 | *OAS1* | 12:112906962 | 1.71 | $9.1 \times 10^{-43}$ | 1.63 | $5.3 \times 10^{-19}$ |
| ENSG00000133106 | *EPSTI1* | 13:42886388 | 1.86 | $3.2 \times 10^{-42}$ | 1.87 | $1.1 \times 10^{-16}$ |
| ENSG00000111335 | *OAS2* | 12:112978519 | 1.44 | $9.9 \times 10^{-41}$ | 1.43 | $6.5 \times 10^{-17}$ |
| ENSG00000187608 | *ISG15* | 1:1013497 | 2.24 | $1.4 \times 10^{-40}$ | 2.43 | $8.2 \times 10^{-18}$ |
| ENSG00000185745 | *IFIT1* | 10:89392623 | 2.38 | $2.7 \times 10^{-39}$ | 2.32 | $9.3 \times 10^{-16}$ |
| ENSG00000157601 | *MX1* | 21:41426239 | 1.9 | $4.8 \times 10^{-39}$ | 1.94 | $4.3 \times 10^{-16}$ |
| ENSG00000137628 | *DDX60* | 4:168216,294 | 1.31 | $6.3 \times 10^{-39}$ | 1.27 | $4.6 \times 10^{-14}$ |

$\beta$ represents DNAm difference between healthy controls and SS patients
Log2FC is the logarithm in base2 of the fold change in gene expression between SS patients and healthy controls.
P is the P-value obtained in the linear regression model adjusted by age, sex, batch effects, and estimated cell proportions.
Genomic positions are based on the hg38 human reference sequence build (GRCh38). TSS=Transcription Start Site

**Table 2.** Genetic association of SS-meQTL and SS-eQTL in SS and other related SADs mediated by DNA methylation or gene expression changes

| Genetic effects on SS mediated by changes in DNA methylation |  |  |  |  |  |  |  |  |  |  |  |
| --- | --- | --- | --- | --- | --- | --- | --- | --- | --- | --- | --- |
| CpG | gene | rsID | SNP Position (hg38 Chr:bp) | A1/A2 | $\beta_{\text{meQTL}}$ | $P_{\text{meQTL}}$ | $\beta_{\text{EWAS}}$ | $P_{\text{EWAS}}$ | OR | P | PRECISESADS |
| cg08099136 | PSMB8 | rs9275569 | 6:32710259 | C/T | -0.018 | $1.92 \times 10^{-04}$ | -0.062 | $2.43 \times 10^{-09}$ | 2.03 | $1.57 \times 10^{-08}$ | SLE (1e-04) |
| cg06033320 | PDE7A | rs16932436 | 8:66036171 | T/C | -0.048 | $2.25 \times 10^{-04}$ | -0.091 | $3.17 \times 10^{-09}$ | 1.76 | 0.0064 | |
| cg14864167 | PDE7A | rs16932436 | 8:66036171 | T/C | -0.069 | $1.83 \times 10^{-05}$ | -0.126 | $2.55 \times 10^{-10}$ | 1.76 | 0.0064 | |
| cg14880222 | IFI44 | rs1051047* | 1:78663909 | A/G | 0.027 | $5.94 \times 10^{-08}$ | -0.036 | $3.61 \times 10^{-09}$ | 0.54 | 0.0127 | SLE (0.042) |
| cg23352030 | PRIC285 | rs76364377* | 20:63687831 | G/A | 0.042 | $1.01 \times 10^{-05}$ | 0.075 | $9.29 \times 10^{-11}$ | 1.60 | 0.0206 | |
| cg06188083 | IFIT3 | rs6583692 | 10:90167339 | A/C | -0.029 | $4.07 \times 10^{-04}$ | -0.150 | $8.85 \times 10^{-18}$ | 1.34 | 0.0236 | |
| cg03879629 | intergenic (CCR cluster) | rs9838739* | 3:46095597 | T/C | -0.046 | $5.21 \times 10^{-05}$ | -0.071 | $7.81 \times 10^{-09}$ | 1.71 | 0.0242 | SLE (0.002), MCTD (0.008), UCTD (0.004) |
| cg12013713 | PARP12 | rs6962291*# | 7:139971218 | T/A | -0.017 | $2.96 \times 10^{-05}$ | -0.048 | $8.54 \times 10^{-09}$ | 1.32 | 0.0270 | |
| cg09858955 | VRK2 | rs11125727 | 2:57698654 | G/T | 0.021 | $2.99 \times 10^{-04}$ | -0.107 | $2.14 \times 10^{-14}$ | 0.78 | 0.0357 | |
| cg12439472 | EPSTI1 | rs4245318 | 13:42984963 | A/C | 0.069 | $2.05 \times 10^{-09}$ | -0.237 | $2.32 \times 10^{-17}$ | 0.77 | 0.0411 | |
| cg03763873 | EPSTI1 | rs4245318 | 13:42984963 | A/C | 0.026 | $6.06 \times 10^{-16}$ | -0.051 | $6.43 \times 10^{-11}$ | 0.77 | 0.0411 | |
| Genetic effects on SS mediated by changes in gene expression |  |  |  |  |  |  |  |  |  |  |  |
| Transcript | gene | rsID | SNP Position (hg38 Chr:bp) | A1/A2 | $\beta_{\text{eQTL}}$ | $P_{\text{eQTL}}$ | $\beta_{\text{diffexp}}$ | $P_{\text{diffexp}}$ | OR | P | PRECISESADS |
| :NSG00000166278 | C2 | rs1054684\$# | 6:32809044 | T/C | 0.300 | $3.57 \times 10^{-04}$ | 0.911 | $1.46 \times 10^{-13}$ | 4.04 | $2.16 \times 10^{-13}$ | SLE(1e-05), MCTD (0.03), UCTD (0.048) |
| :NSG00000204267 | TAP2 | rs4947258 | 6:32779625 | A/G | 0.168 | $3.51 \times 10^{-04}$ | 0.478 | $2.31 \times 10^{-17}$ | 3.53 | $2.90 \times 10^{-12}$ | SLE(1e-05) |
| :NSG00000240065 | PSMB9 | rs3117106 | 6:32375492 | T/C | 0.139 | $2.19 \times 10^{-04}$ | 0.287 | $1.12 \times 10^{-08}$ | 2.22 | $1.23 \times 10^{-07}$ | SLE (0.001) |
| :NSG00000204713 | TRIM27 | rs2523425* | 6:29526364 | C/T | -0.069 | $9.24 \times 10^{-06}$ | -0.136 | $1.06 \times 10^{-06}$ | 1.51 | $7.32 \times 10^{-04}$ | SADsnoSS (0.03) |
| :NSG00000128604 | *IRF5* | rs4731531\$* | 7:128922493 | G/A | 0.084 | $3.45 \times 10^{-04}$ | 0.192 | $6.48 \times 10^{-07}$ | 1.46 | 0.0014 | SLE (0.005) |
| :NSG00000174444 | *RPL4* | rs4482223 | 15:65828392 | A/G | 0.101 | $3.18 \times 10^{-04}$ | -0.195 | $1.01 \times 10^{-09}$ | 0.53 | 0.0019 | RA (0.004) |
| :NSG00000124508 | *BTN2A2* | rs3799378*# | 6:26404046 | A/G | 0.096 | $2.74 \times 10^{-04}$ | 0.228 | $4.61 \times 10^{-08}$ | 1.54 | 0.0025 | |
| :NSG00000234127 | *TRIM26* | rs9261518 | 6:30149108 | G/A | -0.113 | $2.66 \times 10^{-05}$ | 0.154 | $3.65 \times 10^{-09}$ | 0.60 | 0.0029 | UCTD (0.01) |
| :NSG00000126351 | *THRA* | rs12941497* | 17:40099241 | G/A | 0.103 | $2.51 \times 10^{-04}$ | -0.179 | $8.48 \times 10^{-07}$ | 0.63 | 0.0030 | |
| :NSG00000224295 | *AC087380.14* | rs11038373 | 11:5620380 | A/C | 0.090 | $2.76 \times 10^{-05}$ | 1.107 | $1.68 \times 10^{-07}$ | 1.42 | 0.0050 | |
| :NSG00000175518 | *UBQLNL* | rs11038373 | 11:5620380 | A/C | 0.155 | $1.78 \times 10^{-09}$ | 0.857 | $3.35 \times 10^{-10}$ | 1.42 | 0.0050 | |
| :NSG00000175970 | *UNC119B* | rs10849822* | 12:120939591 | G/C | -0.066 | $3.28 \times 10^{-04}$ | -0.161 | $2.88 \times 10^{-09}$ | 1.48 | 0.0075 | SLE (0.02) |
| :NSG00000117226 | *GBP3* | rs599599 | 1:89239059 | T/C | 0.226 | $7.31 \times 10^{-07}$ | 0.424 | $1.02 \times 10^{-06}$ | 1.35 | 0.0125 | |
| :NSG00000120539 | *MASTL* | rs2780683* | 10:27305385 | T/G | -0.187 | $1.41 \times 10^{-06}$ | 0.250 | $5.44 \times 10^{-07}$ | 0.55 | 0.0129 | |
| :NSG00000162614 | *NEXN* | rs6660288 | 1:78927603 | G/A | 0.143 | $2.73 \times 10^{-04}$ | 0.685 | $3.02 \times 10^{-14}$ | 1.36 | 0.0156 | MCTD (0.04), UCTD (0.02) |
| :NSG00000136231 | *IGF2BP3* | rs2722394 | 7:24391386 | T/C | -0.119 | $4.46 \times 10^{-06}$ | 0.423 | $1.01 \times 10^{-06}$ | 0.75 | 0.0168 | |
| :NSG00000002549 | *LAP3* | rs57570581 | 4:17705638 | G/A | -0.216 | $2.76 \times 10^{-05}$ | 0.867 | $2.61 \times 10^{-22}$ | 0.75 | 0.0187 | UCTD (0.02) |
| :NSG00000204516 | *MICB* | rs9348883# | 6:32390672 | T/A | -0.174 | $3.12 \times 10^{-04}$ | 0.242 | $1.19 \times 10^{-09}$ | 0.46 | 0.0276 | |
| :NSG00000123610 | *TNFAIP6* | rs6725961* | 2:151338822 | G/A | -0.306 | $9.39 \times 10^{-05}$ | 0.824 | $2.38 \times 10^{-14}$ | 0.67 | 0.0293 | |
| :NSG00000154451 | *GBP5* | rs76397273* | 1:89333021 | A/G | -0.288 | $9.70 \times 10^{-05}$ | 0.701 | $2.54 \times 10^{-13}$ | 0.67 | 0.0301 | SSc (0.007), UCTD (0.03) |
| :NSG00000128335 | *APOL2* | rs5756497 | 22:37048820 | C/A | 0.123 | $2.03 \times 10^{-04}$ | 0.296 | $8.00 \times 10^{-09}$ | 1.32 | 0.0343 | |
| :NSG00000184898 | *RBM43* | rs289953 | 2:151143385 | T/C | 0.183 | $8.55 \times 10^{-09}$ | 0.378 | $5.03 \times 10^{-12}$ | 1.29 | 0.0398 | SLE (0.03), SSc (0.01) |
| :NSG00000146859 | *TMEM140* | rs17231451 | 7:135130773 | T/C | -0.140 | $1.64 \times 10^{-04}$ | 0.427 | $4.94 \times 10^{-13}$ | 0.75 | 0.0400 | |
| :NSG00000166750 | *SLFN5* | rs11080327* | 17:35244427 | G/A | 0.222 | $4.68 \times 10^{-14}$ | 0.255 | $1.11 \times 10^{-06}$ | 1.28 | 0.0401 | PAPs (0.04) |
| :NSG00000162714 | *ZNF496* | rs56277829* | 1:247365329 | T/C | -0.115 | $3.45 \times 10^{-04}$ | 0.435 | $1.50 \times 10^{-14}$ | 0.76 | 0.0476 | |

Alleles represent major allele first, and then the minor allele, which is the allele tested in each analysis.
$\beta_{meQTL}$ represents the DNA methylation change with the increased in dosage of the minor allele.
$P_{meQTL}$ corresponds to the P value from the linear regression model that regresses out the number of minor alleles for a given SNP to DNA methylation levels adjusting by age, sex, batch effects, estimated cell proportions, disease status and first genetic component.
$\beta_{EWAS}$ represents the DNA methylation difference between SS and healthy controls from the epigenome-wide association study together obtained by linear regression model in which DNA methylation levels are regressed out by SS status and adjusted by age, sex, batch effects and estimated cell proportions.
OR represents the Odd Ratio obtained from genetic association testing based on logistic regression modeling which the SS status is regressed out by number of minor alleles for a given SNP adjusted by age, sex, batch effects, estimated cell proportions and first genetic component and its corresponding.
P represents the P-value obtained in the genetic associations
SAD (OR, P) represents the odd ratio and P value obtained in genetic testing for other diseases. RA = Rheumatoid Arthritis, SLE = Systemic Lupus Erythematosus, UCTD= Undifferentiated Connective Tissue Disease, SSc= Systemic Scleroderma, PAPs = Primary anti-phospholipid syndrome. SADSnoSS = All SADs patients excluding SS.
Genomic positions are based on the hg19 human reference sequence build (GRCh37).
\* eQTL reported in GTEx project (https://www.gtexportal.org/home/), in the case of eQTL the same SNP-gene is reported

**Figure 2.**
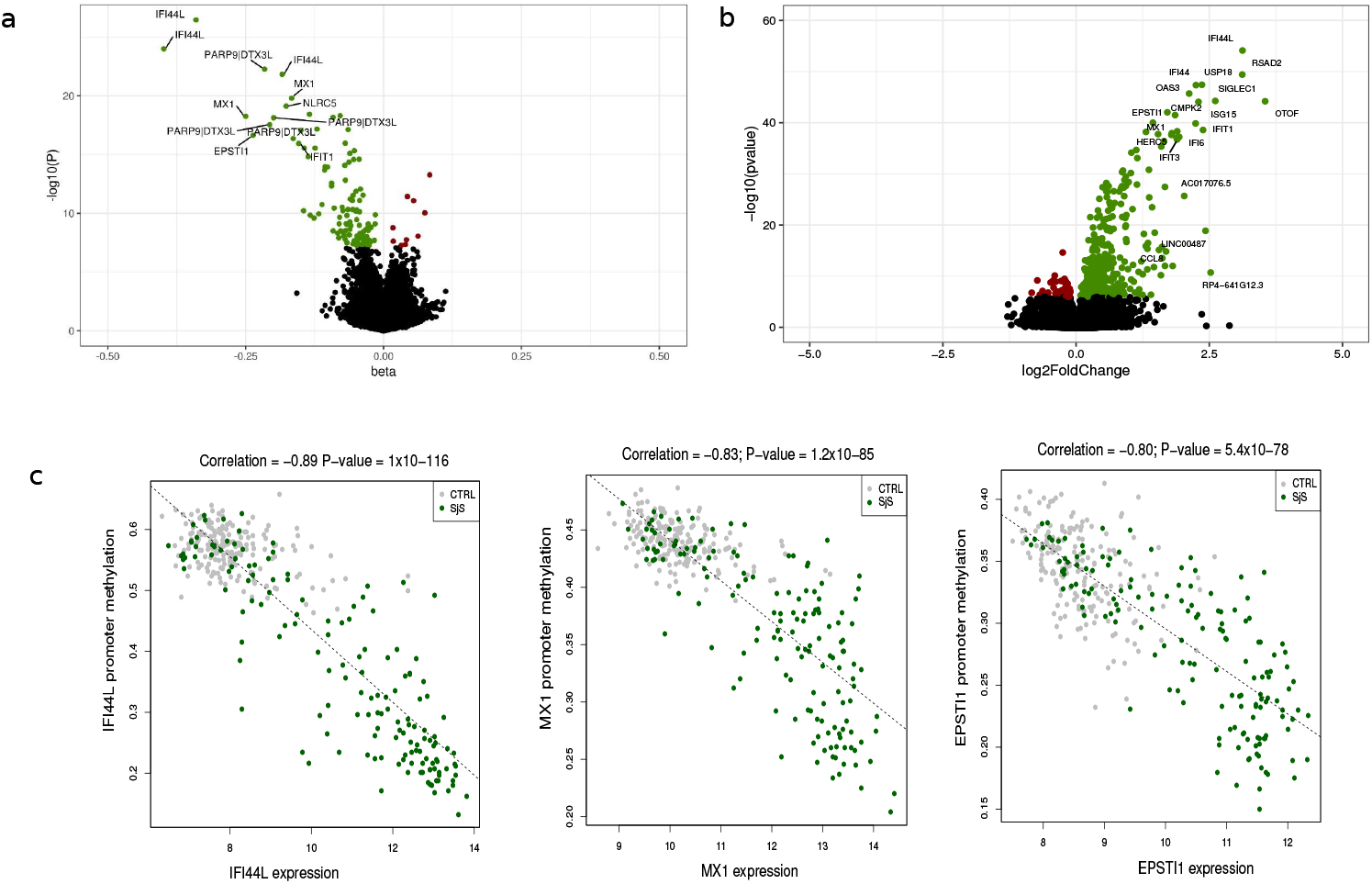
DNA methylation and gene expression patterns associated with SS. Volcano plot for the differential DNA methylation association study in the discovery cohort. P-values are represented on the –log10 scale in the y-axis. The effect size and direction obtained for each CpG site is depicted in the x-axis. Green dots represent significant associations with negative sign (hypomethylation). Red dots represent significant associations with positive signs. The top associations are labeled with gene names. **b**. Volcano plot for the differential expression analysis in the discovery cohort. The effect size and direction obtained for each gene is depicted in the x-axis. Green dots represent significant associations with positive sign (overexpression). Red dots represent significant associations with negative signs. The top associations are labeled with gene names. **c**. Plots showing high correlation between an average of DNA methylation quantified as β-values at the promoters of the most significant SS-associated DMRs and gene expression at the logarithmic scale.

DNA methylation is an epigenetic mark that changes with environmental triggers as for example drugs. We investigated whether our SS-associated signatures were driven by the most common treatments in our SS patients by adjusting the linear model for treatments such as antimalarial, steroids, and immunosuppressive therapy (see details of the treatment applied to the individuals in each cohort in **Supplementary Table 1**). Our results show that most of the SS-DMPs (80.5%) remained significantly associated at our threshold P < 6.4×10^-08^ while the reminder showed suggestive associations, of P <1×10^-04^, indicating that therapy applied to the SS patients does influence the DNA methylation patterns associated with SS only in a minority of sites, with modest effects (**Supplementary Table 3**). In fact, a comparison of the effect size of the 118 significant SS-DMPs obtained in these 2 analyses, when including or not the therapy as covariate in the linear regression, showed high correlation (Pearson’s correlation R= 0.99, P= 2.2×10^-6^). Similar results were also observed in the replication cohort, for which we observed a high replication rate (81.0 %) (**Supplementary Table 4**). Differences in DNA methylation levels and variability of CpGs located in the X-chromosome were also evaluated in females of the discovery cohort, we could not find any significant differences in females at a significance level corrected for multiple testing (data not shown).

**Table 3.** Most significant meQTLs and eQTLs exhibiting disease-dependent genetic effects

| *A. SS-dependent meQTLs* |  |  |  |  |  |  |  |  |  |  |  |
| --- | --- | --- | --- | --- | --- | --- | --- | --- | --- | --- | --- |
| CpG | Gene | SNP | SNP Position (hg38 Chr:bp) | Discovery cohort |  |  |  |  |  | Replication cohort |  |
| | | | | $\beta_{INT}$ | $P_{INT}$ | $\beta_{SS.meQTL}$ | $P_{SS.meQTL}$ | $\beta_{CTRL.meQTL}$ | $P_{CTRL.meQTL}$ | $\beta_{INT}$ | $P_{INT}$ |
| cg08818207 | TAP1 | rs113547322 | 6:32238742 | 0.058 | $1.5 \times 10^{-05}$ | 0.048 | 0.0004 | NA | > 0.05 | 0.059 | 0.0091 |
| cg14392283 | LY6E | rs13273708 | 8:143002764 | -0.018 | $1.6 \times 10^{-04}$ | -0.016 | 0.0006 | NA | > 0.05 | -0.020 | 0.0061 |
| cg01309328 | PSMB8 | rs3134951 | 6:32147308 | -0.040 | $4.0 \times 10^{-04}$ | -0.024 | 0.0124 | NA | > 0.05 | -0.040 | 0.0023 |
| cg14951497 | STAT1 | rs4853645 | 2:191839218 | -0.029 | $5.5 \times 10^{-04}$ | -0.024 | 0.0012 | NA | > 0.05 | -0.038 | 0.0418 |
| cg12906975 | intergenic (LY6E) | rs55937049 | 8:143033566 | -0.013 | 0.0018 | -0.011 | 0.0087 | NA | > 0.05 | -0.017 | 0.0410 |
| cg23387863 | SGK269 | rs1079396 | 15:78137720 | -0.024 | 0.0022 | -0.017 | 0.0119 | NA | > 0.05 | -0.028 | 0.0141 |
| cg10734665 | ATP10A | rs7169481 | 15:25712123 | -0.022 | 0.0031 | 0.018 | 0.0047 | NA | > 0.05 | 0.030 | 0.0138 |
| cg08099136 | PSMB8 | rs3129943 | 6:32370868 | -0.030 | 0.0038 | -0.027 | 0.0050 | NA | > 0.05 | -0.035 | 0.0108 |
| *B.SS-dependent eQTLs* |  |  |  |  |  |  |  |  |  |  |  |
| Gene ID | Gene | SNP | SNP Position (hg38 Chr:bp) | Discovery cohort |  |  |  |  |  | Replication cohort |  |
| | | | | $\beta_{INT}$ | $P_{INT}$ | $\beta_{SS.eQTL}$ | $P_{SS.eQTL}$ | $\beta_{CTRL.eQTL}$ | $P_{CTRL.eQTL}$ | $\beta_{INT}$ | $P_{INT}$ |
| ENSG00000183486 | MX2 | rs9305702 | 21:40755922 | -0.300 | $1.8 \times 10^{-04}$ | -0.302 | $1.8 \times 10^{-04}$ | NA | > 0.05 | -0.315 | 0.0172 |
| ENSG00000198785 | GRIN3A | rs2417310 | 9:101964703 | 0.251 | $4.5 \times 10^{-04}$ | 0.235 | $7.0 \times 10^{-04}$ | NA | > 0.05 | 0.262 | 0.0092 |
| ENSG00000185885 | IFITM1 | rs12364973 | 11:1116140 | 0.386 | $6.9 \times 10^{-04}$ | 0.280 | 0.0130 | NA | > 0.05 | 0.430 | 0.0476 |
| ENSG00000013374 | NUB1 | rs77466830 | 7:151831985 | -0.143 | $7.7 \times 10^{-04}$ | -0.151 | $3.5 \times 10^{-04}$ | NA | > 0.05 | -0.158 | 0.0345 |
| ENSG00000133106 | EPSTI1 | rs9525846 | 13:43816621 | -0.607 | $8.8 \times 10^{-04}$ | -0.577 | $2.4 \times 10^{-03}$ | NA | > 0.05 | -0.651 | 0.0199 |
| ENSG00000188313 | PLSCR1 | rs56077428 | 3:146981084 | 0.471 | 0.0010 | 0.357 | 0.0208 | NA | > 0.05 | 0.600 | 0.0185 |
| ENSG00000157601 | MX1 | rs9305702 | 21:40755922 | -0.602 | 0.0012 | -0.496 | 0.0083 | NA | > 0.05 | -0.779 | 0.0085 |
| ENSG00000247317 | LY6E-DT | rs902834 | 8:142111410 | 0.167 | 0.0038 | 0.129 | 0.0149 | NA | > 0.05 | 0.149 | 0.0469 |
| ENSG00000108691 | CCL2 | rs1490922 | 17:33725536 | 0.351 | 0.0044 | 0.307 | 0.0159 | NA | > 0.05 | 0.367 | 0.0252 |
| ENSG00000187210 | GCNT1 | rs2377425 | 9:75947610 | 0.132 | 0.0047 | 0.083 | 0.0452 | NA | > 0.05 | 0.210 | 0.0068 |

$\beta_{INT}$ represents the interaction effect between a given SNP and SS status in DNA methylation level
$P_{INT}$ represents the P-value obtained for the $\beta_{INT}$ in a linear regression model that adjust for adjusting by SNP, SS status, age, sex, batch effects, estimated cell proportions and the first principal genetic component
$\beta_{SS.meQTL}$ represents the DNA methylation change with the increased in dosage of the minor allele in SS population for a given SNP
$P_{SS.meQTL}$ corresponds to the P value from the linear regression model that regresses out the number of minor alleles for a given SNP to DNA methylation levels adjusting by age, sex, batch effects, estimated cell proportions, disease status and first genetic component in SS population
$\beta_{CTRL.meQTL}$ represents the DNA methylation change with the increased in dosage of the minor allele in the healthy control population for a given SNP
$P_{CTRL.meQTL}$ corresponds to the P value from the linear regression model that regresses out the number of minor alleles for a given SNP to DNA methylation levels adjusting by age, sex, batch effects, estimated cell proportions, disease status and first genetic component in the healthy control population
NA represents effects that are not significant ( $P > 0.05$ )

### Differentially methylated regions associated with SS

We sought to identify differentially methylated regions (DMRs) associated with SS with the purpose of finding subtle, but consistent, methylation changes within a region that could not be detected when analyzing CpG sites by themselves. At FDR 5% threshold, we identified DMRs within 135 gene bodies, 335 promoters and 219 in CpG islands (CGI) (**Supplementary Table 5**). We found that many of the SS-associated DMPs (55.6 %) were SS-associated DMRs, as for examples differentially methylated promoters for *IFI44L* **(Supplementary Fig. 3)**, *OAS2, RSAD2, BST2, PARP9-DTX3L, IFITM1, MX1, EPSTI1* or *LY6E*, and differentially methylated gene bodies of *AGRN, IRF7, B2M, HERC5, ADAR* and *SP100*, among others.

**Figure 3.**
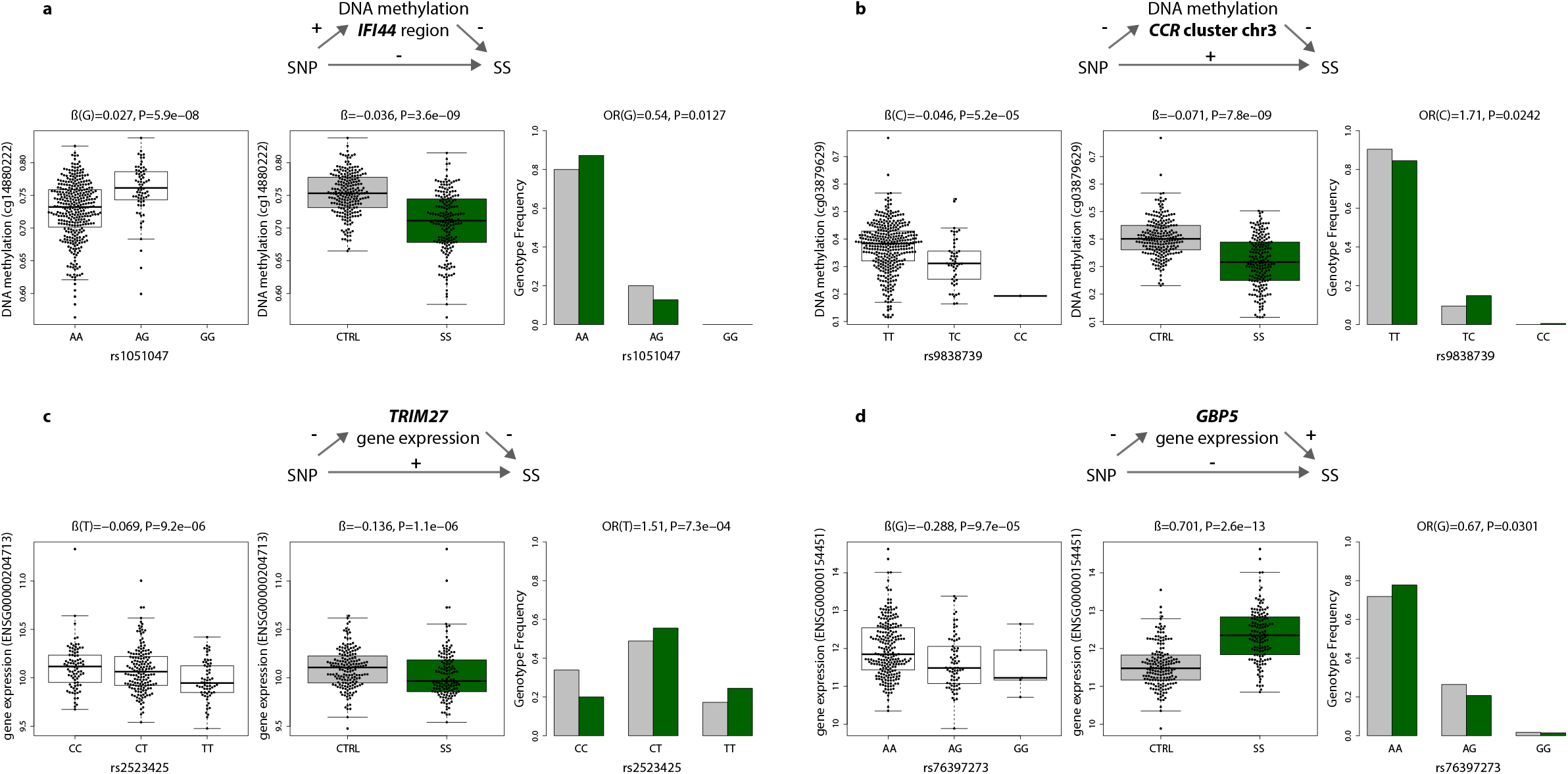
Intermediary role of DNA methylation and gene expression in SS genetic risk. **a**. The minor G-allele of SNP rs1051047 exerts a protective role on SS susceptibility by increasing DNA methylation levels at the upstream region of gene *IFI44* (cg1488022). **b**. The minor C-allele of SNP rs9838739 exerts risk on SS susceptibility by decreasing DNA methylation levels at the intergenic region within the *CCR* cluster in chromosome 3 (cg03879629). **c**. The minor T-allele of SNP rs2523425 exerts risk on SS by decreasing *TRIM27* gene expression. **d**. The minor G-allele of SNP rs76397273 exerts a protective effect on SS by decreasing *GBP5* gene expression. Green boxplots and barplots represent SS population, while grey plots represent the healthy control population. DNA methylation is quantified with β-values, gene expression is at the logarithmic scale.

Importantly, we identified 442 novel differentially methylated genes exhibiting DMRs but for which we did not find DMPs. We could not perform a strict replication of these signatures because DNA methylation was measured in a different array (450K) in the replication sample, and this implies having lower number of CpG sites measured in the pre-defined regions analyzed. However, we found out that up to 45% novel DMRs, lying within 199 genes, showed robust signals in the replication sample (P < 0.05), and were, therefore, validated (**Supplementary Table 5**). Among the novel genes implicated in SS we found new interferon-regulated genes, such as *SAMHD1, ISG15* and *XAF1*; and other proteins related with the immune system such the Tumor Necrosis Factor, *TNF*, the TNF-receptor *CD27*, the chemokine receptor like protein 2, *CCRL2*, and the tyrosine kinase *LCK* (**Supplementary Fig**.**4**). We also discovered a group of genes belonging to the HOX family, such as *HOXB2, HOXD8, HOXA9, HOXA10* or *HOXA4*, that is implicated in transcriptional processes, as well as other transcription factors such *CEBPD* and *GATA2*. Interestingly, we discovered a large group of genes with differential methylated regions implicated in collagen metabolism such as *COL11A2, COL18A1, COL27A1, COL13A1, COL23A1* and *COL5A1* that were not previously identified. In addition, many DMRs, especially those that fell in CGI, were located within long non-coding RNAs (LncRNAs), as for example *RP11-723C11*.*2, RP11-326C3*.*7, RP11-480D4*.*2* or *RP11-89K21*.*1* (see **Supplementary Table 5**). We performed functional enrichment analyses for the set of genes showing lower DNA methylation in the SS group (64%) and separately those with increased DNA methylation (34%). One the hand, DMRs with negative signs showed an enrichment in well known pathways implicated in SS such as interferon and cytokine signaling (R-HSA-913531 and R-HSA-1280215, respectively, from Reactome database), as well as in NOD-like receptor signaling pathway (KEGG path:hsa04621), viral carcinogenesis (KEGG path:hsa05203), transcriptional misregulation in cancer (path:hsa05202), necroptosis (KEEF path:hsa04217), neutrophil degranulation (Reactome R-HSA-6798695) and inflammasomes (Reactome R-HSA-622312), among others. On the other hand, those genes showing DMRs with positive effects were found to be enriched in pathways such as Protein digestion and absorption (path:hsa04974), Collagen biosynthesis and modifying enzymes (Reactome, R-HSA-1650814) and extracellular matrix organization (Reactome R-HSA-1474244) and Vitamin digestion and absorption (KEEG path:hsa04977) **(Supplementary Table 6)**, which represent novel molecular pathways implicated in SS, and support an scenario in which hypermethylation events are also relevant epigenetic changes implicated in SS pathogenesis.

### Differential expression around SS-associated epigenetic signals

In order to detect coordinated epigenetic and transcriptional changes associated with SS, we explored the possibility that the DNA methylation observed at the majority of SS-associated DMRs correlates with the expression of genes in the proximity. We identified 422 differentially expressed genes (DEGs) associated with SS when comparing RNA-seq gene expression data available for 174 SS patients and 135 healthy subjects in our discovery cohort (P < 1.6 x 10^-6^), the majority of them show increased expression in SS patients and were successfully replicated in our independent sample (85.5 % and 88.2%, respectively) (**Fig. 2b and Supplementary Table 7**). *IFI44L* was observed as the most significant differentially expressed gene (Log_2_FC= 3.12; P= 7.6 ×10^-55^; FDR= 3.3×10^-50^) (**Table 1B**). Not surprisingly, many overexpressed genes were related with the IFN signaling (65/422), such as *EPSTI1, RSDA2, USP18 or CMPK2* (**Table 2**), but a large proportion of SS-DEGs belong to other functional categories, being enriched in pathways such as response to external biotic stimulus (GO:0043207), response to stress (GO:0006950), interspecies interaction between organisms (GO:0044419), protein binding (GO:0005515), RNA binding (GO:0003723) or negative regulation of biological processes (GO:0048519) (**Supplementary Table 8)**.

Next, we investigated whether DNA methylation at DMRs and gene expression at DEGs are correlated and if this correlation is associated with SS status and could, therefore, represent coordinated effects on SS, by expression quantitative trait methylation (eQTM) analyses. We observed a total of 48 eQTMs or genes showing a strong correlation between DNA methylation levels and gene expression (Pearson’s coefficient > 0.60, P < 1.6×10^-34^) (**Supplementary Table 9**). Many of the SS-eQTMs fell within the promoters of the IFN-related genes, such as *IFI44L* (**Fig. 2c**), *EPSTI1, MX1*, *DTX3L, PAPR9, LY6E, IFITM3, DDX60, RSAD2, PLSCR1* and *ADAR*. For all of them we observed that decreased DNA methylation strongly correlated with increased expression levels in SS patients, while no correlation was apparent in the healthy population. Interestingly, we observed strong coordinated effects at other non IFN-regulated genes implicated in the immune system such as *CD3D, FGR, PILRA* or *NLRC4*, and in genes with unrecognized function in SS, such as *MDGA1, PM20D1* and *SIRPB2* (Pearson’s coefficient > 0.80). Genes showing correlated patterns of DNA methylation and gene expression were enriched mainly in immune-related gene ontologies being the most significant, immune response (GO:0006955), defense response (GO:0006952), activation of the immune response (GO:0002253) and response to stress (GO:0006950) **(Supplementary Table 10)**. Our results suggest that SS-associated changes in DNA methylation can impact the transcriptional landscape in SS patients, and this could ultimately lead to alteration in the cellular function and immune response.

### Genetic drivers of SS-associated differential methylation and expression

To obtain insight into the extent to which differential methylation and expression associated with SS is genetically controlled, we performed *cis*-meQTL and *cis*-eQTL analyses (see **Methods**). By means of linear regression models that adjust for disease conditions, we found evidence for genetic control in 52% of SS-associated DMPs and 39% of SS-associated DEGs (gene variants no farther than 1Mb to CpG or to transcription start site - TSS). Specifically, at an FDR of 5% we found a total of 4,305 significant meQTLs that included 61 SS-DMPs and 3,508 single-nucleotide polymorphisms (SNPs). We were able to assess the replication of 1475 meQTLs that involved 31 CpGs included in the 450K array and replicated results for 20 CpGs (64%) that were involved in 754 meQTLs (P < 0.05) (**Supplementary Table 11)**. The most significant meQTLs regulate DNA methylation at the *EPSTI1* gene, at an intergenic region in chromosome 3 where CCR cluster is located, and at *VRK2, ADAR, IRF7* and *MX1* genes. On the other hand, we identified a total of 11,399 significant eQTLs, that included 172 SS-DEGs and 10,620 SNPs, from which we could replicate 6414 eQTLs (56%) at a significance threshold of P < 0.05 and with consistent direction of effect, implicating 81 genes (**Supplementary Table 12)**. Genetic variants located close to the TSS of *GBP3* show the strongest association with *GBP3* gene expression, followed by *cis*-genetic variation regulating gene expression of *C3AR1* or *MASTL, ETV7* and *IFITM3*.

### Intermediary role of DNA methylation and gene expression in genetic risk of SS

Then, we investigated the possibility that changes in DNA methylation and gene expression mediate genetic risk in SS, as it has been shown to occur for other autoimmune diseases ^12,29^. For that, we interrogated whether SNPs involved in meQTLs and eQTLs could be linked to the disease and showed allele frequency differences in a set of 391 SS patients and 549 healthy controls in a direction that is consistent with a mediation role of DNA methylation or gene expression (**Methods**). At a Bonferroni-corrected significance of P < 0.0008, we detected risk variants in the HLA region of chromosome 6 that regulate DNA methylation at gene *PSMB8* **(Table 2)**. In this case, the minor T-allele of SNP rs7769693, at the *HLA-DRB9* pseudogene, is associated with increased SS risk and with decreased methylation levels, suggesting that the variant might exert its risk by hypomethylating the *PSMB8* gene. Similar scenarios were observed for genetic variants outside chromosome 6 that show significant association with SS (P < 0.05) and regulate DNA methylation in the downstream region of *IFI44, IFIT3*, at an intergenic position in chromosome 3 within the CCR genetic cluster, and at *PARP12, VRK2, EPSTI1* and *PRIC285* genes **(Table 2, Supplementary Table 13)**. Regarding eQTL results, at a Bonferroni-corrected significant level of P < 0.0002 we detected genetic variants associated with gene expression and SS at three differentially expressed genes *C2, TAP2 and PSMB9* **(Table 3)**. Other genetic variants also show statistical evidence of risk for SS (P < 0.05) and impact gene expression levels at genes such as *TRIM27, IRF5, RPL4, BTN2A2, TRIM26* and *THRA*, among others **(Table3, Supplementary Table 14)**.

To give further support to the functional and genetic link between meQTL and eQTLs with SS and with autoimmune processes, we interrogated whether or not the novel identified SS-risk variants show evidence of association with other SADs for which we had access to genotypic data: systemic lupus erythematosus (SLE), rheumatoid arthritis (RA), systemic sclerosis (SSc), mixed connective tissue disease (MCTD), undifferentiated connective tissue disease (UCTD) and primary antiphospholipid syndrome (PAPs) (**Table 2, Supplementary Table 15**). Moreover, we searched for additional evidence of their functional role in public databases of eQTL and GWAS studies. Importantly, genetic variants regulating DNA methylation at the intergenic region of the CCR cluster in chr3 **(Fig. 3a**) and DNA methylation at the downstream region of *IFI44* gene **(Fig. 3b)** show convincing evidence of their link with other SADS and have been described as eQTL for the same genes (**Supplementary Table 16**). Likewise, genetic variants regulating gene expression of the genes *TRIM27* **(Fig. 3c)**, *BTN2A2, UNC119B, GBP5* **(Fig. 3d)** and *SLFN5* show significant associations with other SADS. A link with gene expression has been previously observed in eQTL studies. Altogether, these results support the scenario in which -associated differences in DNA methylation and gene expression are intermediates of genetic risk for SS, and provides a framework for the discovery of new risk *loci* associated with SS via alteration of regulatory landscapes.

### Disease-dependent genetic effects on SS-associated differential methylation and gene expression

Finally, we hypothesized that there might exist genetic variants associated with SS risk whose effect on molecular phenotypes depends on the specific environment of altered immune activation originated during the disease process. In order to identify such disease-dependent genetic effects or *disease-interacting QTLs*, we performed a gene-by-environment interaction meQTLs and eQTLs analyses in which we looked for gene variants that interact with disease status on shaping DNA methylation and gene expression levels at SS-associated DMPs and DEGs. We considered relevant and robust disease interacting QTLs those that accomplish the following strict conditions: i) an interaction effect that passes a significance threshold of P_INTER_ < 0.005 in the discovery cohort and a significance threshold of P_INTER_ < 0.05 in the replication cohort, with consistent direction of the effect, ii) genetic variant associated with DNA methylation or gene expression only in the SS group (P_SS.QTL_ < 0.05) and without evidence of genetic associations in the healthy population (P_CTRL.QTL_ > 0.05), iii) genetic variants that show a minor allele frequency higher than 0.10 in the SS group.

For DNA methylation, we observed convincing evidence for disease-interacting meQTLs at 8 SS-associated DMPs within genes *TAP1, LY6E, PSMB8, STAT1, SGK269* and *ATP10A* (**Table 3** and **Supplementary Table 17**). The most significant gene-environment interaction (ß_INTER_ = -0.018, P_INTER_ = 2.0×10^-04^) that fell outside the HLA region involved the genetic variant rs13273708 at chromosome 8 and DNA methylation at *LY6E* (cg14392283) (**Fig. 4a, Table 3**). For this variant, the minor C-allele is associated with a decrease in *LY6E*-DNA methylation only in SS patients, but this association is not observed in the healthy population (**Table 3**). A similar scenario is observed for the *STAT1* gene for which we found the second most significant disease-dependent meQTLs effect (ß_INTER_ = -0.029, P_INTER_ = 5.47×10^-04^) outside the HLA region (**Fig. 4b**). Another convincing example implicates the interaction between SS status and rs1079396 in shaping DNA methylation levels at *SGK269* gene (ß_INTER_ = -0.024, P_INTER_ = 0.003, **Fig. 3c**), and the interaction with rs7169481 in shaping DNA methylation at *ATP10A* (cg10734665, ß_INTER_ = 0.022, P_INTER_ = 0.003, **Fig. 4d**). For gene expression, some IFN-inducible genes such as *MX2, IFITM1, EPST1, MX1* and *LY6E-DT* show convincing evidence that their expression is genetically regulated in a disease-specific manner (**Table 3** and **Supplementary Table 18**). The most significant disease-dependent effect was observed for *MX1* gene expression (ß_INTER_ = -0.300, P_INTER_ =1.8×10^-0^, **Fig. 4e**), which is overexpressed in SS patients, and only genetically regulated within the disease population. In this case, the minor T-allele of rs9305702 is associated with decreased *MX1* gene expression in SS patients, without evidence of association in the healthy population (**Fig. 4e**). Other genes such as *IFITM1* (**Fig. 4f**), *CCL2* (**Fig. 4g**), *NUB1* (**Fig. 4h**), *PLSCR1, GRIN3A* and *GCNT1* also show disease-specific eQTLs that regulate their SS-associated differential expression **(Table 3**).

**Figure 4.**
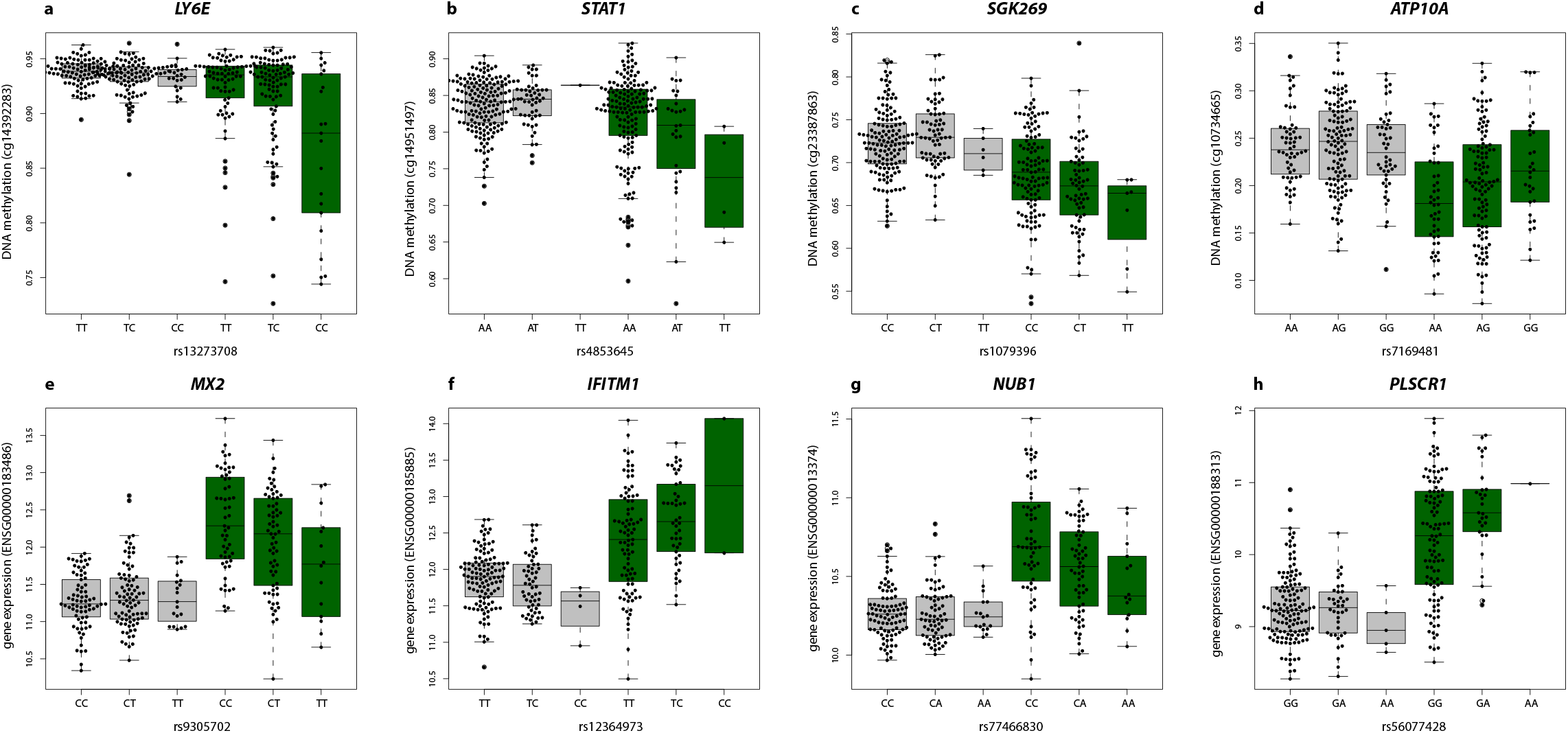
Disease interacting QTLs. **a**. The minor C-allele of SNP rs13273708 is associated with a decrease in DNA methylation levels at *LY6E* gene only in SS patients (ß_SS.meQTL_ =-0.018, P_SS.meQTL_ = 6×10^-04^), but not in the healthy population (P_SS.meQTL_ > 0.05). **b**. The minor A-allele of SNP rs902834 decreases the DNA methylation level at *STAT1* only in SS patients (ß_SS.meQTL_ =-0.024, P_SS.meQTL_ = 0.0012), and not the healthy population (P_CTRL.meQTL_ > 0.05). **c**. The minor T-allele of rs1079396 is associated with *SGK269*-methylation in SS patients (ß_SS.meQTL_ =-0.017, P_SS.meQTL_ = 0.0119), but not in the healthy population (P_CTRL.meQTL_ > 0.05). **d**. The minor G-allele of rs7169481 in *ATP10A* is associated with increased DNA methylation at *ATP10A* in SS patients (ß_SS.meQTL_ = 0.018, P_SS.meQTL_ = 0.0047). However, in the healthy population this allele has no significant effect (P_CTRL.meQTL_ > 0.05). **e**. The minor T-allele of the rs9305702 genetic variant is associated with a decreased *MX2* gene expression in SS patients (ß_SS.eQTL_ = -0.302, P_SS.eQTL_ =1.8×10^-04^), and shows no evidence of association in the healthy population (P_CTRL.eQTL_ > 0.05). **f**. The minor C-allele of rs12364973 is associated with an increased *IFITM1* gene expression in SS patients (ß_SS.eQTL_ =0.28, P_SS.eQTL_ =0.012) and shows no evidence of association in the healthy population (P_CTRL.eQTL_ > 0.05). **g**. In SS patients, *NUBI* expression decreases with the dose of the minor A-allele of rs77466830 (ß_SS.eQTL_ = -0.15, P_SS.eQTL_ = 3.5×10^-04^); however, in the healthy population it remains stable (P_CTRL.eQTL_ > 0.05). **h**. In SS patients, *PLSCR1* expression increased with the dose of the minor A-allele of rs56077428 (ß_SS.eQTL_ = 0.357, P_SS.eQTL_ = 0.0208); however, in the healthy population it remains stable (P_CTRL.eQTL_ > 0.05). Green boxplots represent SS population, while grey boxplots represent the healthy control population. DNA methylation is quantified with β-values, gene expression is at the logarithmic scale.

For all those disease-interacting meQTLs and eQTLs, the same trend is observed in the replication sample (**Supplementary Table 17 and 18**) and we found no evidence for them to be genetic regulators of DNA methylation and/or gene expression in a model that corrects for disease status and includes the whole population (**Supplementary Table 17 and 18**), or in other studies that have interrogated healthy populations **(Supplementary Table 19)** ^8,10,30^. Our findings reveal a differential genetic architecture of gene regulation between SS patients and the healthy population, and suggest that the specific immunological and pathological conditions in autoimmunity modify how genes regulate the transcriptional landscapes of immune cells, which likely has an impact in the function and fate of cells.

## DISCUSSION

Here, we report for the first time a comprehensive integrative large-scale analysis of DNA methylation, gene expression and genetic data in a large SS cohort. To the best of our knowledge, this is the first time that the EPIC array has been used to assay the differential methylation between SS patients and healthy control samples, which has allowed us to interrogate the methylation status of more than 750.000 CpG sites across the genome. Our findings have confirmed the well-known overall hypomethylated state, and also the overexpression, of many IFN-related genes in SS patients as it was previously described in different tissues and cell types ^18,19,21,22,31,32^. We observed that the majority of SS-associated DMPs exhibited higher DNA methylation variability in SS patients when compared to healthy controls. Increased in epigenetic variability is also observed for other autoimmune conditions ^29,33,34^ and is correlated with disease progression, severity and resistance to therapy ^35^. Epigenetic plasticity allows a rapid adaptation of immune cells to external conditions as effective defense against pathogens but it might also reflect regulation of DNA methylation by diverse transcription factors involved in different inflammatory and immune signaling during disease progression ^35,36.^.

Applying statistical methods based on gene set enrichments allowed us to detect a large number of novel genes associated with SS that exhibit DNA methylation differences of smaller magnitude but with consistent epigenetic patterns along pre-defined functional regions. We found differentially methylated regions at genes related to IFN-pathways that could not have been identified when analyzing individual CpG sites, such as those found in *SAMHD1, ISG15* and *XAF1* genes, and also in other immune-related genes such as the chemokine *CCRL2* or the T-cell tyrosine kinase *LCK*. Importantly, a high percentage of DMRs are hypermethylated in SS patients, a trend that had not been observed in EWAS that analyze individual CpG sites, where most of the signals identified correspond to hypomethylation events. Functional analyses in the group of genes exhibiting positive effects revealed an enrichment in important pathways such as those related to the metabolism of collagen and/or implicated in extracellular matrix organization that have not been previously detected, but could explain the increased degradation of extracellular matrix structures and the significant loss of collagen observed in the lacrimal gland and other tissues in SS, and, therefore, have a key role in its pathogenesis^37^. We anticipate that future efforts in biomarker discovery will successfully recognize the SS-associated hypermethylated signals of good utility for SS diagnosis and for discrimination with other related diseases.

When exploring the relationship between DNA methylation at DMR and gene expression profiles of nearby genes, we found that many IFN-regulated genes, such as *IFI44L, RSAD2, OAS2, EPSTI1, PARP9, CMPK2 or MX1*, and also other immune-related genes like *NLRC4, CD28, FGR* and *S100A9*, show strong correlation between DNA methylation and gene expression. This pattern suggests that there is a coordinated regulation of these molecular marks that is dependent on the disease status and suggests that disease driven-hypomethylation could result in enhanced transcriptional activity of nearby regions, and be potential novel targets for drug development.

In this study, we discovered new *loci* associated with SS whose functional mechanisms could be the alteration of epigenetic states or gene expression profiles. The association that we have found between genetics, DNA methylation and SS in the *CCR* gene cluster in chromosome 3 is very interesting and represents a novel genetic risk variant. Our study shows that the minor C-allele of rs9838739, located in an active regulatory region overlapping many transcription factor binding sites and upstream the *CCR* gene cluster, increases risk for SS and several related systemic autoimmune diseases (SLE, MTD and UCTD) and decreases DNA methylation levels at this dense regulatory region. Previous eQTL studies from GTEx project identified that C-allele is associated with lower *CCR5* expression in whole blood^30^. Interestingly, the lack of *CCR5* on dendritic cells of a NOD mouse, an experimental model for SS disease and diabetes, promotes a proinflammatory environment in submandibular glands, a target and affected tissue in SS patients ^38^. Furthermore, *CCR5* expression is decreased on circulating monocytes from SS patients and is correlated with increased levels of inflammatory chemokines ^39^. Our results, together with previous findings, suggest that genetically determined reduced methylation at *CCR* cluste*r*-methylation, could contribute to SS presumably via enhanced gene expression of inflammatory chemokines.

Our integrative approach has also revealed that *IRF5* is a *locus* implicated in SS, which likely impacts the disease through gene expression changes, as it was described for SLE ^40^. Other than this, we identified other putative novel genetic determinants of SS with regulatory function that have been independently replicated in related systemic autoimmune diseases, as for example those regulating the expression of *TRIM27, TRIM26, RLP4, NEXN, LAP3, GBP5, RBM43* and *SLNF5* genes. For example, we found that the T-minor allele of SNP rs2523425 on its upstream intergenic region, is associated with decreased *TRIM27* expression, as it has been already reported ^30^, and with increased odds for patients with SS of being diagnosed with other systemic autoimmune diseases. We corroborate in this work that *TRIM27* is down-regulated in SS patients, as it has been previously reported ^41^. TRIM27 is a molecule that inhibits the innate immune response and has been recently shown to negatively regulate NOD2 mediated signaling by physical interaction and degradation^42^. These data suggest that genetically determined reduced *TRIM27* expression in SS patients can lead to enhanced innate responses via abnormally enhanced NOD2 activity.

Importantly, the findings of this work corroborate our hypothesis that genetic variants may only manifest genetic regulatory effects in the specific context of altered immune activity, in this case exhibited in SS patients. We successfully identified and replicated a number of SS-dependent meQTLs that regulate DNA methylation at *STAT1, LY6E* and *ATP10A* genes, among others. STAT1 is a signal transducer and activator of transcription that is activated in response to several cytokines as IFN-alpha, IFN-gamma or IL-6. Stimulation of monocytes and B-cells in SS patients leads to increased sensitivity of immune cells from SS patients to STAT1-activating signals that might partly explain the IFN signature observed in SS ^43^. In line with these results, our transcriptional data shows overexpression of *STAT1* in SS patients. On the other hand, *LY6E*, an IFN-regulated gene that encodes for the Lymphocyte Antigen 6E, is hypomethylated and overexpressed in SS patients. Interestingly, a previous study discovered trans-eQTL effects on *LY6E* gene expression that is dependent on immune activation^44^, supporting that its genetic regulation is context-specific. An earlier GWAS study on cytokine responses found that genetic variants at *ATP10A* are associated with IFN-gamma production in response to vaccinia virus in subjects who had received the smallpox vaccine ^45^. These findings, and our results, support a scenario in which inter-individual genetic variation at this gene impacts IFN-regulated gene expression only upon immune activation.

Some of the genes whose expression is genetically regulated in a disease-specific manner belong to the group of IFN-regulated genes, such as *MX1, MX2, EPST1* and *IFITM1*, but we also discovered other genes with immune-related function such as *CCL2* and *PSLCR1. CCL2* encodes for a chemokine ligand that is secreted during inflammatory processes whose serum circulating levels are increased in SS ^46^. Intriguingly, the expression of *CCL2* is genetically regulated upon *in vitro* stimulation but it shows no genetic regulation in naïve conditions ^47^. *PSLCR1*, the Phospholipid scramblase 1 gene, has been found to be differentially methylated and expressed in SS, SLE and MCTD ^21,48,49^. It is a DNA-binding transcriptional activator that amplifies the IFN-mediated antiviral response and induces cytokine expression and cell proliferation. Interestingly, a previous study has shown that the genetic regulation of *PSLCR1* expression is stronger upon IFN stimulation ^44^. We also found disease-interacting genetic variants whose implication in systemic autoimmunity has not been well described. For example, *NUB1* is an IFN-inducible gene that downregulates NEDD8, an ubiquitin-like protein. *NUB1* overexpression is known to induce inhibition of cell growth in cancer cells ^50^. Its implication in autoimmune processes is uncertain but it could be related to lymphoma development.

To wrap up, this study confirmed the whole blood epigenetic and transcriptional IFN signature previously observed in SS patients. We discovered novel genes exhibiting differentially methylated regions and identified important novel pathways involved in the disease, as of example those related to collagen metabolism and extracellular matrix organization, that are characterized by hypermethylation events. By integrating multiple omics layers of information, we identified a group immune-related dysregulated genes exhibiting coordinated epigenetic and transcriptional changes associated with SS. Finally, we expanded the list of genetic variants with regulatory function that are involved in SS pathogenesis, many of which only show a functional effect in the specific environmental context of immune activation that is characteristic of an autoimmune state. Our study serves to discover new *loci* and pathways involved in SS, to recognize the importance of hypermethylation events in SS pathogenesis, and it contributes to unravel the genetic regulatory architecture of Sjögren’s Syndrome, which hopefully will motivate research for the discovery of new drugs and biomarkers for SS in the near future.

## MATERIAL AND METHODS

### Participant recruitment

Samples included in this study were recruited from the PRECISESADS study, which is a European multi center, non-randomized, and observational clinical study with recruitment performed between December 2014 and December 2018 at 19 institutions in 9 countries. PRECISESADS included patients affected by systemic autoimmune (around 400 by disease or group of diseases: rheumatoid arthritis, scleroderma or systemic sclerosis, primary Sjögren’s syndrome, systemic lupus erythematosus, and primary antiphospholipid syndrome, mixed connective tissue disease and undifferentiated connective tissue disease) and 554 healthy controls ^51^. An ethical protocol was prepared, reached consensus across all partners, academic and industrial, translated into all participant’s languages and approved by each of the local ethical committees of the clinical recruitment centers, and all experimental protocols were approved by each of the local committees. For a list of local committees and centers involved in PRECISESADS please see **Supplementary Note1**. All patients recruited to the study were aged 18 years or older and signed an informed consent form, and all methods were carried out in accordance with relevant guidelines and regulations. The study adhered to the standards set by International Conference on Harmonization and Good Clinical Practice, and to the ethical principles that have their origin in the Declaration of Helsinki (2013). The protection of the confidentiality of records that could identify the included individuals is ensured as defined by the EU Directive 2001/20/EC and the applicable national and international requirements relating to data protection in each participating country.

A total of 558 samples were included in the core analyses of the present study **(Fig. 1)**, that included SS patients and healthy controls for which DNA methylation and genotype data was available. All patients diagnosed with SS (N=278) fulfilled the diagnostic criteria of the American and European community published in 2002 ^52^. The majority of the SS patients (94.8%) were females with a mean of age of 58.8 ± 12.4 years. Healthy individuals (N=280), i.e., not having any history of autoimmune or infectious diseases, were included as controls, and matched to cases to the extent possible. The 74.4 % of the healthy subjects were females with a mean of age of 45.1 ± 13.3 years **(Supplementary Table 1)**. Samples were divided in two sets; a discovery cohort formed by 189 SS patients and 220 healthy subjects and a replication cohort that included 60 SS patients and 89 healthy individuals. Epidemiological and clinical features of the samples included in the discovery and the replication cohorts are summarized in **Supplementary Table 1**.

### Genome-wide DNA methylation data and differential analyses

DNA methylation data of the discovery cohort was obtained using the Infinium Methylation EPIC BeadChip (Illumina, San Diego, CA, USA) that covers more than 800,000 CpG sites. For replication cohort, we had also available genome-wide DNA methylation data profiled obtained by Infinium Methylation 450K BeadChip (Illumina, San Diego, CA, USA), which covers more than 400,000 CpG sites, most of them included in the EPIC array.

After DNA extraction from whole peripheral blood and bisulfite conversion, the genome for each sample was amplified, fragmented and hybridized to the corresponding Illumina arrays according to the manufacturer’s protocol. The quality control of samples and the normalization of the data was performed using the meffil R software ^53^. Samples were excluded based on the detection P criteria > 99%, poor bisulfite conversion based on control dashboard check, and sex mismatches according to failed chromosome X and Y clustering. Probes were filtered out based on detection P > 0.01 in > 95% of samples. Additionally, probes located at the X and Y chromosomes were separated in different datasets to avoid gender bias. Probes with genetic variants at their CpG sites were also excluded. After applying these filtering steps we obtained 776,284 and 433,337 autosomic probes in the discovery dataset and in the replication dataset, respectively. A total of 17,530 probes located in the X chromosome passed the QC in the discovery cohort and 9,282 in the replication cohort. DNA methylation was measured as a beta value ranging from 0 to 1. Zero represents an unmethylated state (0% molecules methylated at a particular sites) while 1 represents a fully methylated state (100% molecules methylated). After QC, the raw methylation beta values were background corrected and normalized using the functional normalization within the meffil R-package.

Differentially DNA methylated positions (DMPs) associated with SS were identified in our discovery cohort using a linear regression model that regresses out the SS status on DNA methylation levels at each CpG. The model was adjusted by sex, age, the first genetic principal component, the observed blood cell proportions obtained at the time of sample extraction, and batch effects as covariates. The technical variables Sentrix_ID and Sample_Plate corrected the batch effect. Blood cell proportions were obtained using Duraclone tubes (Beckman Coulter) by whole blood flow cytometry, as described in Jamin et al. ^54^ (see **Supplementary Note2** for a list of investigators involved in the PRECISESADs Flow Cytometry Study Group). They were calculated as percentages of total leukocytes. Given the population stratification observed in our data, the first principal component was included in the linear model as covariate. We also searched for variable methylated positions (VMP) that show DNA methylated variance differences between SS patients and healthy samples. For that, we first obtain the residuals of DNA methylation levels after correcting for all covariates in a linear regression model. Then, we searched for variance differences in DNA methylation residuals between cases and controls by applying a Levene’s test that accounts for mean differences. A Bonferroni-corrected threshold of P < 6.4 x 10^-08^ was established to consider DMPs and VMPs as significant. For each significant DMP the mean DNA methylation in cases and controls was calculated. The independent cohort was used to evaluate the robustness of the significant associations. The same steps were followed in the replication set. In this case, the replication significance threshold was established at P < 0.05. All statistical analyses were performed using R (v3.4.2) ^55^. The effect of treatment on the epigenetic associations found was evaluated. First, treatments were included as covariates in the linear model and we compared the results of the treatment-adjusted and treatment unadjusted models. Specifically, we controlled for the use of antimalarial, steroids and immunosuppressants, since we observed that more than 10 SS patients had these drugs prescribed. Then, stratified analyses according to the presence/absence of these treatments were performed. The differences of the DNA methylation between non-treated or treated cases and controls was evaluated using a linear regression model adjusted by sex, age, the first genetic principal component, the cell proportions, and batch effects. **Supplementary Table 1** shows the treatments applied to the SS patients during the time of the study.

Differential DNA methylation regions (DMRs) associated with SS were estimated using mCSEA (methylated CpGs Set Enrichment Analysis) R package that implements a Gene Set Enrichment Analysis method (GSEA) to identify DMRs ^56^. Briefly, the first step of mCSEA consists in ranking all the CpG probes by differential methylation and subsequently evaluates the enrichment of CpG sites belonging to the same region in the top positions of the ranked list by applying the GSEA implementation of the *fgsea* package. *mCSEA* allows to perform analysis based on promoters, gene bodies, and CGIs that are defined based on R annotation packages form Illumina for 450K and EPIC arrays, respectively. A DMRs was considered as significant with a threshold FDR < 0.05 and with a minimum number of 5 CpGs associated to the region.

We used the mCSEA R package ^56^ to perform expression quantitative trait methylation (eQTM) analysis to discover significant associations between these DMRs and an expression alteration in the closer genes. In this case, the leading edge CpGs of each region is first defined and averaged for each region in each sample. Then, Pearson’s correlation coefficient is calculated between each region’s methylation and the proximal genes expression within 1500 base pairs upstream and downstream from the region.

### RNA-seq data and differential expression analysis

We analysed gene expression data available from 179 SS patients and 247 healthy subjects. RNA-seq data was obtained as 50 bp single end reads using an Illumina HiSeq 2500 sequencer and its proprietary base caller. Fastq data was aligned against human genome GRCh19 using STAR aligner^57^ with GENCODE v19 [URL: https://www.gencodegenes.org/releases/19.html] annotation as single pass alignment. Read counts were obtained using RSEM ^58^. Samples were included only if they had more than 8 million reads, a RIN score > 8 and did not appear as outliers in a principal component analysis. After filtering, a total of 42,878 genes were included in the study.

Differential expression analysis was performed using a linear model that regressed out SS status on read-counts as gene expression levels adjusted by age, sex, the first genetic principal component, the cell proportions and sequencing pool (POOL) and RNA integrity number (RIN) as batch effects. A Bonferroni-adjusted P-value threshold < 1.16 ×10^-6^ was established as significant to identify the differentially expressed genes (DEGs) in a discovery cohort formed by 135 SS patients and 174 healthy controls. In the replication cohort (44 SS patients and 73 healthy controls), the same steps were followed while a replication significance threshold was established at P< 0.05. The analyses were performed using DESeq2 ^59^

### Genotype profiling

Genomic DNA from whole blood was obtained by standard methods. The samples were genotyped using HumanCore -12-v1-0-B, InfiniumCoreExome-24v1-2 and InfiniumCoreExome-24v1-3, all of them of Illumina (San Diego, CA, USA). Only genetic variants, not indels, present on all three platforms were considered for data cleaning and analysis. Quality controls (QC) were performed using PLINK v1.9 ^60^. Genetic markers were removed if these had a call rate < 90%, exhibited a significant differential missingness between cases and controls (P< 1×10^-4^), and showed a significant deviation from Hardy-Weinberg equilibrium (P< 0.01 in controls and P< 1×10^-4^ in cases). Variants with a minor allele frequency of less than 5% were excluded from the analysis. Samples were excluded of the study if they had a call rate < 95% and also high heterozygosity rate, i.e., they deviated 6 standard deviations from the centroid. Duplicated or related individuals were identified using identity-by-descent criteria with REAP ^61^. Samples were excluded applying a threshold of kinship coefficient < 0.25. Samples with less than 55 % European ancestry were considered outliers and excluded from the analyses. Finally, a total of 218,947 variants passed data filtering.

Inference methods based on linkage disequilibrium structure was used in order to increase the number of genetic markers. Imputation was performed using the Michigan Imputation Server [URL: https://imputationserver.sph.umich.edu/index.html] ^62^ and Haplotype Reference Consortium (HRC) as reference panel [URL: http://www.haplotype-reference-consortium.org/]^63^. We considered the imputed genotypes with a info-value (Minimac R^2^) higher than 0.7, i.e., 70% of reliability. Imputed variants were also filtered according to the protocol described above.

In order to study the population structure and prevent population stratification the individual admixture frequency for each individual was estimated. In addition, population stratification was also analyzed by principal component analysis (PCA). Ancestry per cent for each individual was estimated using FRAPPE ^64^, and a set of 2,707 independent genetic variants that maximized the differences between populations and clustering by K=5, i. e, the 5 global populations: American, African, South Asian, East Asian and European. European published populations from 1000 Genomes phase 3 [URL: http://www.internationalgenome.org/category/phase-3/] were included as reference panel. PCAs were also calculated using SMARTPCA from the Eigensoft software^65^ and the same independent set of markers. In this analysis, only the European population from 1000 Genomes, excluding Finns, was included as reference population of our samples^66^. Six standard deviations from the centroid were used as a threshold for data filtering.

### Quantitative trait loci analysis and genetic associations

Genetic regulation of DNA methylation and gene expression was explored by the quantitative trait *locus* (QTL) approach using Matrix eQTL^67^. A linear regression model was performed to evaluate the effect of closely located SNPs (no further than 1Mb to CpG site or TSS) with a MAF > 0.05 on DNA methylation levels of DMPs and on gene expression patterns of DEGs. In this case, the gene expression data were firstly normalized into variance stabilizing transformations (VST) using DESeq2^59^. The linear model was corrected by age, sex, disease status, the first genetic principal component, cell proportions, and the respective batch effect. A P-value threshold of false discovery rate (FDR) < 0.05 was considered as significant.

To investigate the possibility of different behavior of genetic effects between SS patients and healthy subjects in regulating DNA methylation and gene expression, meQTL and eQTL interacting analyses that included an interaction term between the considered molecular phenotype and the disease status was included in the linear regression model. Because of the lower statistical power of this analysis, we investigated only genetic variants reaching a minimum allele frequency of 10% among SS patients. We considered as significant those interaction signals that pass a suggestive P<0.005 in the discovery cohort, that show consistent direction of effects in the replication sample and reached a P< 0.05 that could only be detected as meQTL or eQTL among SS patients (P< 0.05), but not in healthy subjects (P> 0.05).

Case-control genetic association analyses were conducted using PLINK v1.9 ^60^. Logistic regressions under the allelic additive model were performed to interrogate the association between SS diagnosis and the dosage of the minor allele in SNPs involved in meQTL and eQTLs correcting for the first genetic principal component. We used a P < 0.05 to report significant genetic associations. Moreover, we tested the genetic association of SS-associated genetic variants with other SADs comparing the allele frequencies of 261 SS, 314 SLE, 297 SSc, 332 RA, 59 PAPs, 69 MCTD and 111 UCTD patients vs. 457 healthy controls.

### Bioinformatics tools

Significant DMPs were annotated to genes and gene locations according to annotation files provided by Illumina, from which we obtained a list of unique differentially methylated genes. Enrichment analyses based on gene ontology (GO) terms were performed using the web server ConsensusPathDB-human [URL: http://cpdb.molgen.mpg.de/CPDB] ^68^ that integrates different types of functional interactions from 30 public sources in order to assemble a more complete and a less biased picture of cellular biology. Currently, ConsensusPathDB contains metabolic and signaling reactions, physical protein interactions, genetic interactions, gene regulatory interactions and drug-target interactions in human, mouse, and yeast.

## Data Availability

The cohort datasets generated and analyzed during the current study will be available upon request through ELIXIR platform.

## Data Availability

The cohort datasets generated and analyzed during the current study are available upon request through ELIXIR platform.

## ACKNOWLEDGEMENTS

Funding for the preparation of this manuscript has received support from the Innovative Medicines Initiative Joint Undertaking under grant agreement nº 115565, resources composed of the financial contribution from the European Union’s Seventh Framework Program (FP7/2007-2013) and the EFPIA companies’ in kind contribution. MT is supported by a Spanish grant from Health Department, Junta de Andalucía (PI/0017/2016) and through the Innovative Medicines Initiative 2 Joint Undertaking under grant agreement No 806975. This Joint Undertaking receives support from the European Union’s Horizon 2020 research and innovation programme and EFPIA. EC-M was funded by the Postdoctoral Training Subprogramme Juan de la Cierva-Ministry of Economy and Competitiveness (FJCI_2014_20652). Maria Teruel was supported. We thank Ralf Lesche for the production of RNASeq data and Marc Torres Ciuró for design support.

## AUTHOR CONTRIBUTIONS

EC-M, MEA-R, and MT contributed to the conception and design of the study and also drafted the manuscript. The PRECISESADS Clinical Consortium members agreed on the clinical data, recruited the patients, and performed their detailed clinical assessment. PRECISESADS Cytometry Consortium obtained flow cytometry data. MT, EC-M, GB, MB, EP, MK, SZ, AB and MM-B contributed to pre-processing and/or analyzing the data. MEA-R, JM, JO-P and EB contributed to data generation and sample recruitment. MEA-R coordinated the entire PRECISESADS project. All authors read and approved the final manuscript.

## MATERIALS & CORRESPONDENCE

Correspondence and material request should be addressed to Elena Carnero-Montoro and/or to Marta E. Alarcón-Riquelme

### PRECISESADS Flow Cytometry Study Group

Montserrat Alvarez^1^, Damiana Alvarez-Errico^2^, Nancy Azevedo^3^, Nuria Barbarroja^4^, Anne Buttgereit^5^, Qingyu Cheng^6^, Carlo Chizzolini^1^, Jonathan Cremer^7^, Aurélie De Groof^8^, Ellen De Langhe^9^, Julie Ducreux^8^, Aleksandra Dufour^1^, Velia Gerl^6^, Maria Hernandez-Fuentes^10^, Laleh Khodadadi^6^, Katja Kniesch^11^, Tianlu Li6, Chary Lopez-Pedrera8, Zuzanna Makowska^5^, Concepción Marañón^13^, Brian Muchmore^13^, Esmeralda Neves^3^, Bénédicte Rouvière^14^, Quentin Simon^14^, Elena Trombetta^12^, Nieves Varela^13^ & Torsten Witte^11^.

^1^Immunology and Allergy, University Hospital and School of Medicine, Geneva, Switzerland. ^2^Chromatin and Disease Group, Bellvitge Biomedical Research Institute (IDIBELL), Barcelona, Spain. ^3^Serviço de Imunologia EX-CICAP, Centro Hospitalar e Universitário do Porto, Porto, Portugal. ^4^IMIBIC, Reina Sofia Hospital, University of Cordoba, Córdoba, Spain. 9Bayer AG, Berlin, Germany. ^5^Pharmaceuticals Division, Bayer Pharma, Berlin, Germany.^6^Department of Rheumatology and Clinical Immunology, Charité University Hospital, Berlin, Germany. ^7^Laboratory of Clinical Immunology, Department of Microbiology and Immunology, KU Leuven, Leuven, Belgium. ^8^Pôle de Pathologies Rhumatismales Inflammatoires et Systémiques, Institut de Recherche Expérimentale et Clinique, Université Catholique de Louvain, Brussels, Belgium. ^9^Division of Rheumatology, University Hospitals Leuven and Skeletal Biology and Engineering Research Center, KU Leuven, Leuven, Belgium.^10^UCB, Slough, UK. ^11^Klinik für Immunologie Und Rheumatologie, Medical University Hannover, Hannover, Germany. ^12^Laboratorio di Analisi Chimico Cliniche e Microbiologia - Servizio di Citofluorimetria, Fondazione IRCCS Ca’ Granda Ospedale Maggiore Policlinico di Milano, Milan, Italy. ^13^GENYO, Center for Genomics and Oncological Research Pfizer/University of Granada/Andalusian Regional Government, Granada, Spain. ^14^NSERM, UMR1227, CHRU Morvan, Lymphocytes B et Autoimmunité, Univ Brest, BP 824, Brest, France

### PRECISESADS Clinical Consortium

Lorenzo Beretta^1^, Barbara Vigone^1^, Jacques-Olivier Pers^2^, Alain Saraux^2^, Valérie Devauchelle-Pensec^2^, Divi Cornec^2^, Sandrine Jousse-Joulin^2^, Bernard Lauwerys^3^, Julie Ducreux^3^, Anne-Lise Maudoux^3^, Carlos Vasconcelos^4^, Ana Tavares^4^, Esmeralda Neves^4^, Raquel Faria^4^, Mariana Brandão^4^, Ana Campar^4^, António Marinho^4^, Fátima Farinha^4^, Isabel Almeida^4^, Miguel Angel Gonzalez-Gay Mantecón^5^, Ricardo Blanco Alonso^5^, Alfonso Corrales Martínez^5^, Ricard Cervera^6^, Ignasi Rodríguez-Pintó^6^, Gerard Espinosa^6^, Rik Lories^7^, Ellen De Langhe^7^, Nicolas Hunzelmann^8^, Doreen Belz^8^, Torsten Witte^9^, Niklas Baerlecken^9^,Georg Stummvoll^10^, Michael Zauner^10^, Michaela Lehner^10^, Eduardo Collantes^11^, Rafaela Ortega-Castro^11^, Mª Angeles Aguirre-Zamorano^11^, Alejandro Escudero-Contreras^11^, Mª Carmen Castro-Villegas^12^, Norberto Ortego^12^, María Concepción Fernández Roldán^12^, Enrique Raya^13^, Inmaculada Jiménez Moleón^13^, Enrique de Ramon^14^, Isabel Díaz Quintero^14^, Pier Luigi Meroni^15^, Maria Gerosa**Error! Bookmark not defined**., Tommaso Schioppo^15^, Carolina Artusi^15^, Carlo Chizzolini^16^, Aleksandra Zuber^16^, Donatienne Wynar^16^, Laszló Kovács^17^, Attila Balog^17^, Magdolna Deák^17^, Márta Bocskai^17^, Sonja Dulic^17^, Gabriella Kádár^17^,Falk Hiepe^18^, Velia Gerl^18^, Silvia Thiel^18^, Manuel Rodriguez Maresca^19^, Antonio López-Berrio^19^, Rocío Aguilar-Quesada^19^, Héctor Navarro-Linares^19^

^1^Referral Center for Systemic Autoimmune Diseases, Fondazione IRCCS Ca’ Granda Ospedale Maggiore Policlinico di Milano, Italy. ^2^Centre Hospitalier Universitaire de Brest, Hospital de la Cavale Blanche, Brest, France. ^3^Pôle de pathologies rhumatismales systémiques et inflammatoires, Institut de Recherche Expérimentale et Clinique, Université catholique de Louvain, Brussels, Belgium. ^4^Centro Hospitalar do Porto, Portugal. ^5^Servicio Cantabro de Salud, Hospital Universitario Marqués de Valdecilla, Santander, Spain. ^6^Hospital Clinic I Provicia, Institut d’Investigacions Biomèdiques August Pi i Sunyer, Barcelona, Spain. ^7^Katholieke Universiteit Leuven, Belgium. ^8^Klinikum der Universitaet zu Koeln, Cologne, Germany. ^9^Medizinische Hochschule Hannover, Germany. ^10^Medical University Vienna, Vienna, Austria. ^11^Servicio Andaluz de Salud, Hospital Universitario Reina Sofía Córdoba, Spain. ^12^Servicio Andaluz de Salud, Complejo hospitalario Universitario de Granada (Hospital Universitario San Cecilio), Spain. ^13^Servicio Andaluz de Salud, Complejo hospitalario Universitario de Granada (Hospital Virgen de las Nieves), Spain. ^14^Servicio Andaluz de Salud, Hospital Regional Universitario de Málaga, Spain. ^15^Università degli studi di Milano, Milan, Italy. ^16^Hospitaux Universitaires de Genève, Switzerland. ^17^University of Szeged, Szeged, Hungary. ^18^Charite, Berlin, Germany. ^19^Andalusian Public Health System Biobank, Granada, Spain.

