## Supplementary Note1 for "An integrative multi-omics approach in Sjögren’s Syndrome identifies novel genetic drivers with regulatory function and disease-specificity"

**List of Approved Local Ethical Committees:**

• Referral Center for Systemic Autoimmune Diseases, Fondazione IRCCS Ca’ Granda Ospedale Maggiore Policlinico di Milano, Comitato Etico Italy.

• Centre Hospitalier Universitaire de Brest, Hospital de la Cavale Blanche, Avenue Tanguy Prigent 29,609, Brest, France. Comite de Protection des Personnes Ouest VI.

• Pôle de pathologies rhumatismales systémiques et inflammatoires, Institut de Recherche Expérimentale et Clinique, Université catholique de Louvain, Brussels, Belgium. Comité d`Èthique Hospitalo-Facultaire.

• Centro Hospitalar do Porto, Portugal. Comissao de ética para a Saude – CES do CHP.

• Servicio Cantabro de Salud, Hospital Universitario Marqués de Valdecilla, Santander, Spain. Comite ético de investigacion clinical de Cantabria. IDIVAL.

• Hospital Clinic I Provicia, Institut d’Investigacions Biomèdiques August Pi i Sunyer, Barcelona, Spain. Comité Ética de Investigación Clínica del Hospital Clínic de Barcelona. HOSPITAL CLíNIC DE BARCELONA.

• Katholieke Universiteit Leuven, Belgium. Commissie Medische Ethiek UZ KU Leuven /Onderzoek.

• Klinikum der Universitaet zu Koeln, Cologne, Germany. Geschaftsstelle Ethikkommission

• Medizinische Hochschule Hannover, Germany. Ethikkommission.

• Medical University Vienna, Vienna, Austria. Ethik Kommission. Borschkegasse.

• Servicio Andaluz de Salud, Hospital Universitario Reina Sofía Córdoba, Spain. Comité de Ética e la Investigación de Centro de Granada (CEI – Granada).

• Servicio Andaluz de Salud, Complejo hospitalario Universitario de Granada (Hospital Universitario San Cecilio), Spain. Comité de Ética e la Investigación de Centro de Granada (CEI – Granada).

• Servicio Andaluz de Salud, Complejo hospitalario Universitario de Granada (Hospital Virgen de las Nieves), Spain. Comité de Ética e la Investigación de Centro de Granada (CEI – Granada).

• Servicio Andaluz de Salud, Hospital Regional Universitario de Málaga, Spain. Comité de Ética e la Investigación de Centro de Granada (CEI – Granada).

• Università degli studi di Milano, Milan, Italy. Policlinico di Milano, Comitato Etico Italy.

• Hospitaux Universitaires de Genève, Switzerland. DEAS –Commission Cantonale d`´ethique de la recherche Hopitaux universitaires de Geneve.

• University of Szeged, Szeged, Hungary. Csongrad Megyei Kormanyhivatal.

• Charite, Berlin, Germany. Ethikkommission.

• Andalusian Public Health System Biobank, Granada, Spain.

• Comité de Ética e la Investigación de Centro de Granada (CEI – Granada).
