## Supplementary Figures for "An integrative multi-omics approach in Sjögren’s Syndrome identifies novel genetic drivers with regulatory function and disease-specificity"

**Suppl. Figure 1.** Distribution of the SS-associated DMP within genes.

|  | Open Sea | Shelf | Shore | Island |
| --- | --- | --- | --- | --- |
| Promoter | 21 | 5 | 31 | 1 |
| Body | 20 | 6 | 18 | 4 |
| 3'UTR | 3 | 1 | 1 |  |
| Intergenic | 2 | 1 | 4 |  |

**Suppl. Figure 2.** Examples of SS-DMP in the discovery cohort (EPIC) and the replication cohort (450K).

Methylation is quantified with β-values

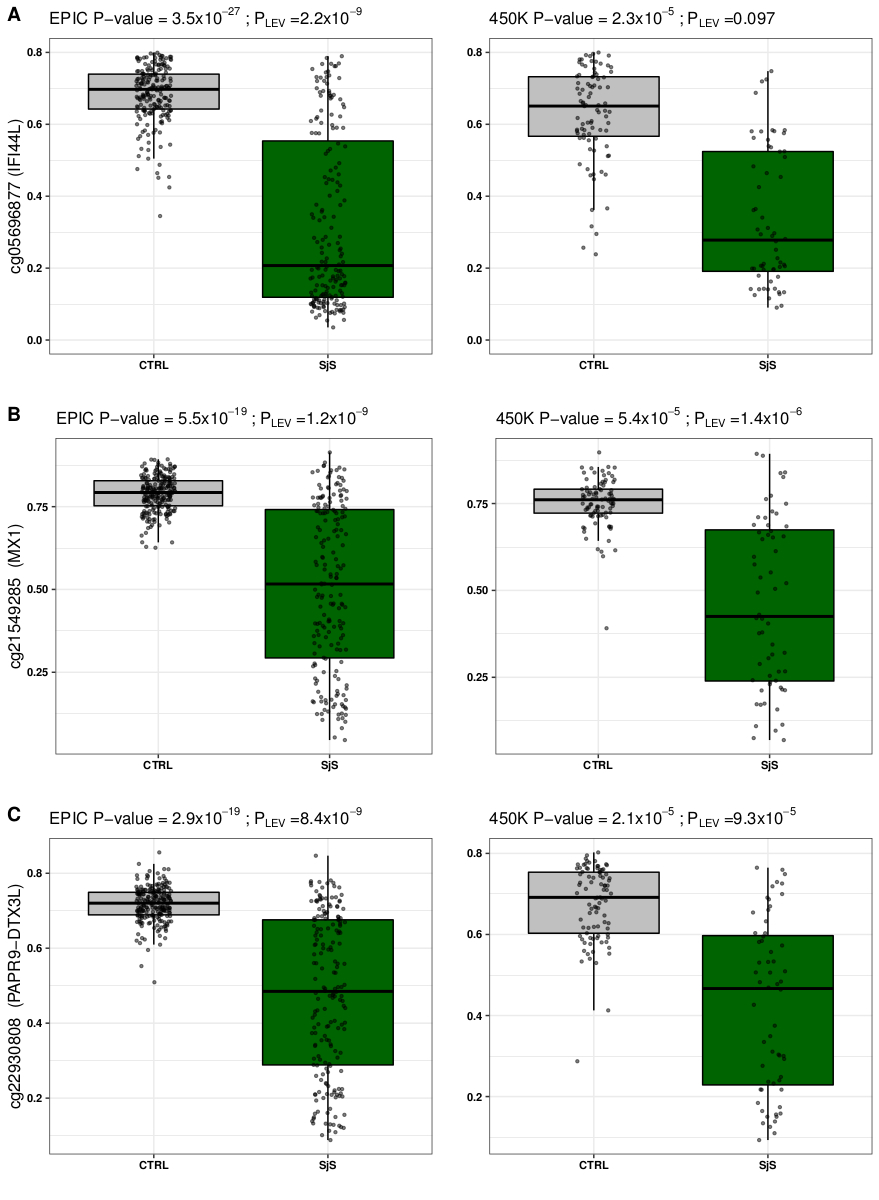

**Suppl. Figure 3.** Genomic context of *IFI44L* promoter.

Methylation is quantified with β-values. Each point represents the methylation of each sample. Lines link the mean methylation of each group. KS leading edge panel marks with black bars those CpGs contributing to the ES and with grey bars the rest of them. This plot was obtained with mCSEAPlot() function, implemented in mCSEA package.

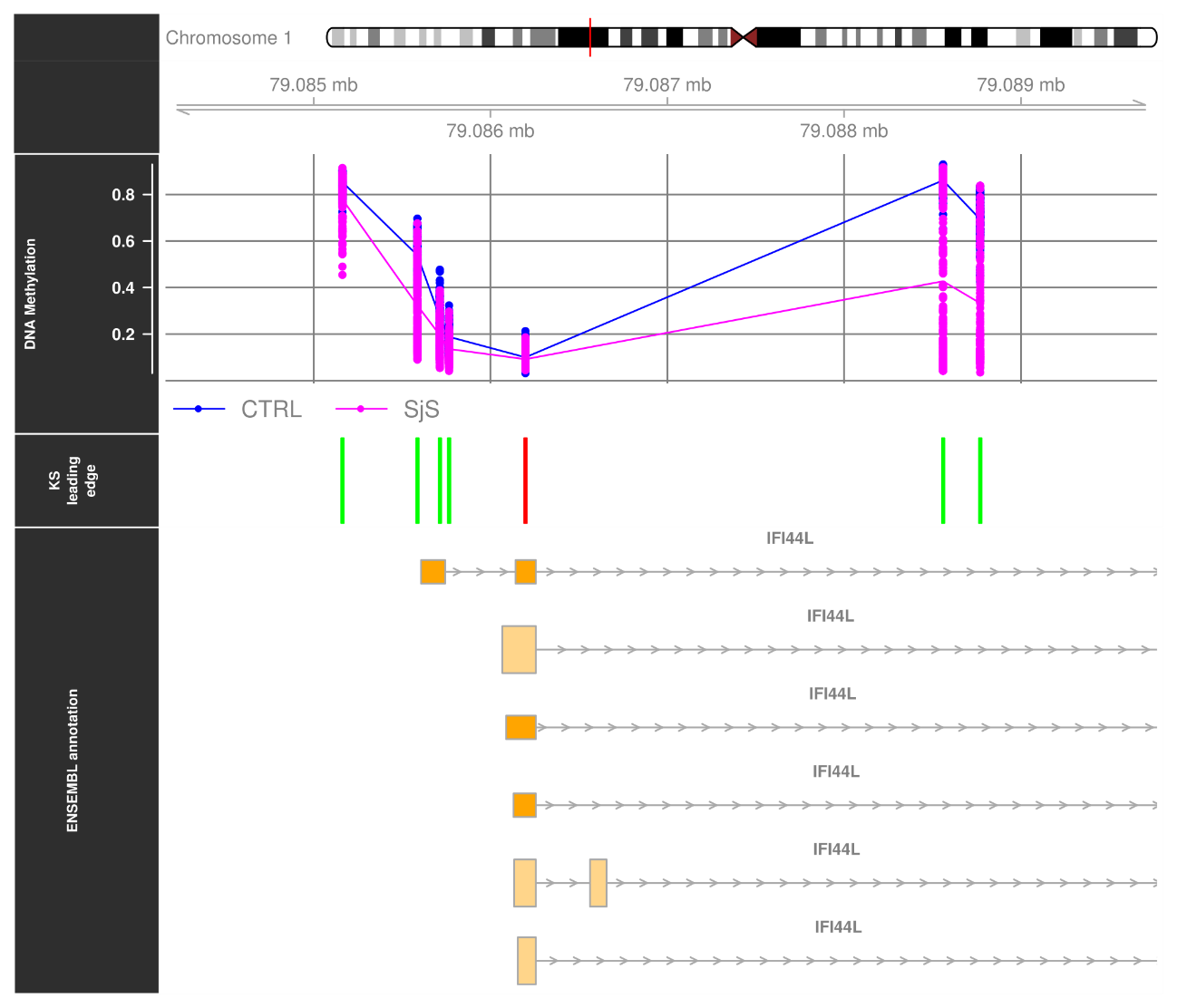

**Suppl. Figure 4.** Genomic context of *LCK* promoter.

Each point represents the methylation of each sample. Lines link the mean methylation of each group. KS leading edge panel marks with black bars those CpGs contributing to the ES and with grey bars the rest of them. This plot was obtained with mCSEAPlot() function, implemented in mCSEA package.

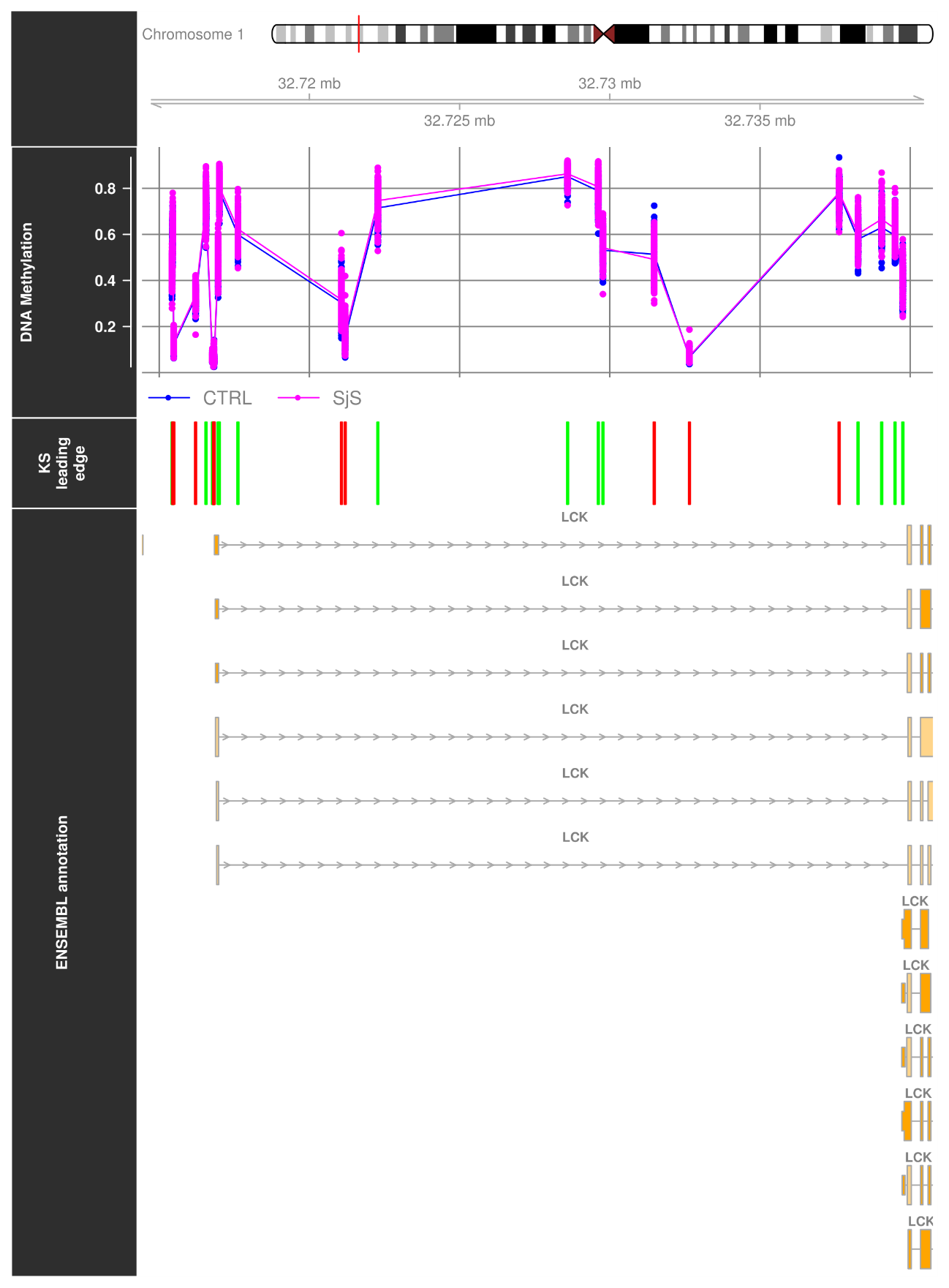
